# Effectiveness and Safety of Glucocorticoids to Treat COVID-19: A Rapid Review and Meta-Analysis

**DOI:** 10.1101/2020.04.17.20064469

**Authors:** Shuya Lu, Qi Zhou, Liping Huang, Qianling Shi, Siya Zhao, Zijun Wang, Weiguo Li, Yuyi Tang, Yanfang Ma, Xufei Luo, Toshio Fukuoka, Hyeong Sik Ahn, Myeong Soo Lee, Zhengxiu Luo, Enmei Liu, Yaolong Chen, Chenyan Zhou, Donghong Peng, on behalf of COVID-19 evidence and recommendations working group

## Abstract

**Background:** Glucocorticoids are widely used in the treatment of various pulmonary inflammatory diseases, but they are also often accompanied by significant adverse reactions. Published guidelines point out that low dose and short duration systemic glucocorticoid therapy may be considered for patients with rapidly progressing COVID-19 while the evidence is still limited.

**Methods:** We comprehensively searched electronic databases and supplemented the screening by conducting a manual search. We included RCTs and cohort studies evaluating the effectiveness and safety of glucocorticoids in children and adults with COVID-19, SARS and MERS, and conducted meta-analyses of the main indicators that were identified in the studies.

**Results:** Our search retrieved 23 studies, including one RCT and 22 cohort studies, with a total of 13,815 patients. In adults with COVID-19, the use of systemic glucocorticoid did not reduce mortality (RR=2.00, 95% CI: 0.69 to 5.75, *I*^*2*^=90.9%) or the duration of lung inflammation (WMD=-1 days, 95% CI: -2.91 to 0.91), while a significant reduction was found in the duration of fever (WMD=-3.23 days, 95% CI: -3.56 to -2.90). In patients with SARS, glucocorticoids also did not reduce the mortality (RR=1.52, 95% CI: 0.89 to 2.60, *I*^*2*^=84.6%), duration of fever (WMD=0.82 days, 95% CI: -2.88 to 4.52, *I*^*2*^=97.9%) or duration of lung inflammation absorption (WMD=0.95 days, 95% CI: -7.57 to 9.48, *I*^*2*^=94.6%). The use of systemic glucocorticoid therapy prolonged the duration of hospital stay in all patients (COVID-19, SARS and MERS).

**Conclusions:** Glucocorticoid therapy was found to reduce the duration of fever, but not mortality, duration of hospitalization or lung inflammation absorption. Long-term use of high-dose glucocorticoids increased the risk of adverse reactions such as coinfections, so routine use of systemic glucocorticoids for patients with COVID-19 cannot be recommend.

## Background

An infectious disease caused by a previously unknown type of coronavirus, the severe acute respiratory syndrome coronavirus 2 (SARS-CoV-2), emerged at the end of December 2019 and has posed a major challenge to the public health worldwide (1). The World Health Organization (WHO) officially named the disease as Corona Virus Disease 2019 (COVID-19) on February 11, 2020 (2). On March 11, 2020, the WHO declared COVID-19 as a global pandemic (3). Globally, as of 2:00am CEST, 12 April 2020, there have been 1,699,595 confirmed cases of COVID-19, including 106,138 deaths, reported to WHO (4).

At present, there are no specific drugs for the prevention and treatment of COVID-19, and symptomatic supportive treatment remains the most effective method of care. Full-genome sequencing and phylogenetic analyses have indicated SARS-CoV-2 is a distinct clade of beta-coronaviruses, related to the Middle East Respiratory Syndrome Coronavirus (MERS-CoV) and Severe Acute Respiratory Syndrome coronavirus (SARS-CoV). Therefore, the management of COVID-19 can benefit from experience from the SARS and MERS epidemics (5). Glucocorticoids were commonly used for the treatment of SARS and MERS especially in critically ill people (6-7), and are also widely used in the treatment of COVID-19.

There are conflicting opinions about the use of glucocorticoids to treat patients with COVID-19. It is suggested that current clinical evidence does not support the use of glucocorticoids, which may cause several side effects (8-9). However, clinicians who are on the front line of the epidemic have proposed that short-term glucocorticoid therapy with small or medium dose could be beneficial for patients with severe conditions (10). The current guidelines on COVID-19 are also inconsistent about the use of glucocorticoids. Some guidelines suggested trying short-term therapy with medium or small doses of glucocorticoids for patients with rapid or severe disease progression, but according to the WHO guidelines glucocorticoids should only be used under clinical trial conditions (11-13). Effective evidence related to glucocorticoids to treat COVID-19 is still lacking.

Therefore, the purpose of this study is to systematically retrieve and summarize the current evidence of the effectiveness and safety of glucocorticoid therapy for patients with COVID-19, aiming to provide the best decision-making basis for the prevention and control of the COVID-19 epidemic.

## Methods

### Search strategy

Two experienced librarians searched the following databases from January 1st, 2003 to March 31th, 2020: The Cochrane library, MEDLINE (via PubMed), Embase, Web of Science, CBM (China Biology Medicine), CNKI (China National Knowledge Infrastructure), and Wanfang Data. We used the following search: (“COVID-19” OR “SARS-CoV-2” OR “2019 novel coronavirus” OR “2019-nCoV” OR “Wuhan coronavirus” OR “novel coronavirus” OR “Wuhan seafood market pneumonia virus” OR “Wuhan virus” OR “MERS” OR “SARS” OR “Severe Acute Respiratory Syndrome” OR “Middle East Respiratory Syndrome Coronavirus” OR “Influenza”) AND (“adrenal cortex hormones” OR “betamethasone valerate “OR “glucocorticoids” OR “methylprednisolone” OR “Cortisone” OR “Dexamethasone” OR “Cortodoxone” OR “Hydrocortisone”). We also searched clinical trial registry platforms (the World Health Organization Clinical Trials Registry Platform, US National Institutes of Health Trials Register and the International Standard Randomized Controlled Trial Number (ISRCTN) Register), Google Scholar (https://scholar.google.nl/) and preprint platforms BioRxiv (https://www.biorxiv.org/), MedRxiv (https://www.medrxiv.org/) and SSRN (https://www.ssrn.com/index.cfm/en/). In addition, we searched the reference lists of the identified systematic reviews to find further potential studies, and supplemented screening Google Scholar by conducting a manual search every day before submission. The search strategy was constructed with the assistance of a specialist in information retrieval (15). The details of the search strategy can be found in the Supplementary Material 1.

### Inclusion and exclusion criteria

We included all studies on glucocorticoid therapy for patients diagnosed with COVID-19, SARS or MERS, without restricting the diagnostic criteria. We included randomized controlled trials (RCTs) and cohort studies comparing glucocorticoid therapy versus placebo or comparing a combination of glucocorticoids and symptomatic treatment with symptomatic treatment alone. The primary outcome of interest was mortality, and secondary outcomes included duration of lung inflammation absorption, duration of hospital stay, duration of fever, and other adverse effects like coinfections (bacterial or fungal infections), kaliopenia, and osteonecrosis of femoral head (ONFH).

We excluded conference abstracts, articles written in languages other than English or Chinese and studies where we could not retrieve the full text or essential data were missing. All the reasons for exclusion of ineligible studies were recorded, and the process of study selection was documented using a PRISMA flow diagram (14).

### Study selection

After eliminating duplicates, two researchers (S Lu and L Huang) independently screened the literature in two steps using the EndNote software. In the first step, all titles and abstracts were screened using pre-defined criteria to exclude irrelevant articles. In the second step, full-texts of the potentially eligible and unclear studies were reviewed to decide about final inclusion. Disagreements were discussed or solved with a third researcher (Q Shi). All the reasons for exclusion of ineligible studies were recorded, and the process of study selection was documented using a PRISMA flow diagram (16).

### Data extraction

Two researchers (S Lu and Q Zhou) independently extracted the data and information from all included studies by using a standardized data collection form. Extracted data included 1) basic information: first author, publication year, and the type of study design; 2) participants: disease, severity of disease, age distribution, and total number of patients; 3) details of the intervention and control groups: type, dosage and treatment course of glucocorticoid therapy; and 4) outcomes: for dichotomous data, we abstracted the number of events and total number of patients per group; and for continuous data, we abstracted the means, standard deviations (SD), and the total number of patients per group. For data that were missing or reported in unusable way, we reported the findings descriptively.

### Risk of bias assessment

Two researchers (S Lu and S Zhao) independently assessed the potential bias in each included study, and discrepancies were resolved by discussion or consulting a third researcher (Q Shi). We assessed the risk of bias in RCTs using the Cochrane risk-of-bias tool (17), which consists of seven domains: random sequence generation, allocation concealment, blinding of participants and personnel, blinding of outcome assessment, incomplete outcome data, selective outcome reporting, and other bias. Each domain was graded as “Low”, “Unclear”, or “High”. For cohort studies, we used the Newcastle-Ottawa Scale (NOS) (18), which contains eight domains: representativeness of exposure cohorts, selection of non-exposure cohorts, determination of exposure, outcome events that did not occur before study initiation, comparability of cohort based on design or analysis, assessment of outcome events, adequacy of follow-up time, and completeness of follow-up.

### Quality of the evidence

Two researchers (Zhou Q and Shi Q) assessed the quality of evidence independently using the Grading of Recommendations Assessment, Development and Evaluation (GRADE) tool (19-20). We produced a “Summary of Findings” table using the GRADEpro software. The quality of evidence can be downgraded based on five factors (study limitations, consistency of effect, imprecision, indirectness and publication bias) and upgraded based on three factors (large magnitude of effect, dose-response relation and plausible confounders or biases) (21-26). The quality of evidence of each outcome is then classified as “high”, “moderate”, “low” or “very low”.

### Data synthesis

We conducted meta-analyses by using Stata 14 software (Stata Corp LLC). For dichotomous data, we calculated risk ratios (RR) with 95% confidence intervals (CI); for continuous data, we calculated weighted mean differences (WMD) with 95% CI. Missing data were dealt with according to the Cochrane Handbook for Systematic Reviews of Interventions (27). As clinical and methodological heterogeneity in study design, characteristics of participants, interventions and outcome measures was expected, we used random-effects models (28). Two-sided *P* values <0.05 were considered statistically significant. Statistical heterogeneity was assessed with the *I*^*2*^ statistic, >50% indicating substantial heterogeneity. If we detected heterogeneity, we performed subgroup analyses by the severity of the disease or the age of patients, and also considered sensitivity analyses where one study was excluded at a time. Egger test was used to assess publication bias (14). Each comparison is presented by the name of the first author and the year of publication.

As COVID-19 is a public health emergency of international concern and the situation is evolving rapidly, our study was not registered in order to speed up the process.

## Results

### Basic characteristics

The rapid review identified 2509 publications, of which 23 studies (one RCT and 22 cohort studies) (29-51) met our inclusion criteria and were included. The literature screening process is shown in *Figure 1*. One study included adult patients with severe MERS, 17 studies included patients with SARS, and the remaining five studies were on patients with COVID-19 (*Table 1*). Due to the insufficient representativeness and follow-up time, only three articles scored higher than six out of nine points. The methodological quality of included cohort studies was poor. The risk of bias of included RCT was unclear because of unclear risks of selection bias, detection bias and reporting bias (*Table 2, Table 3*).

**Table 1.**
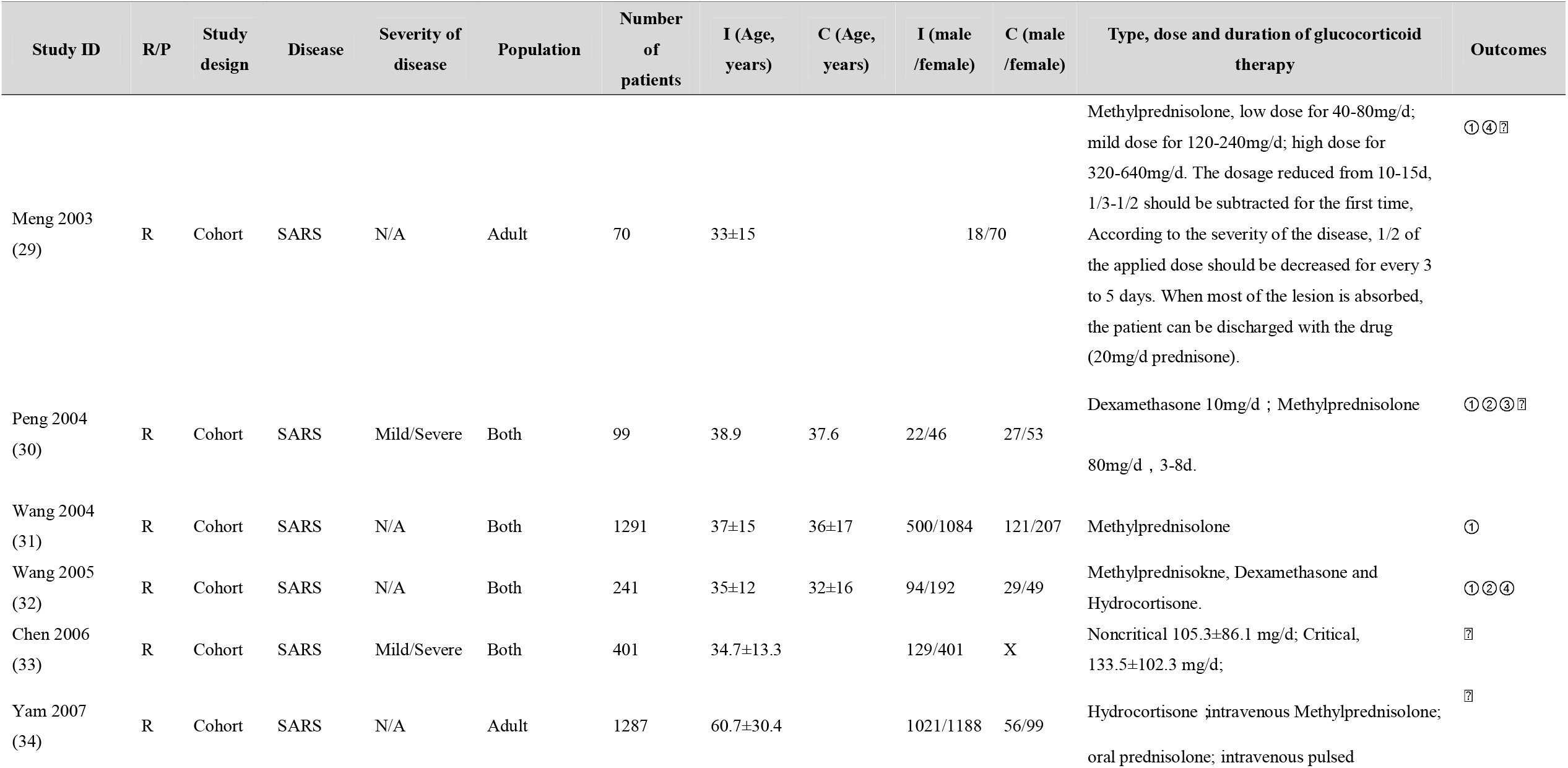
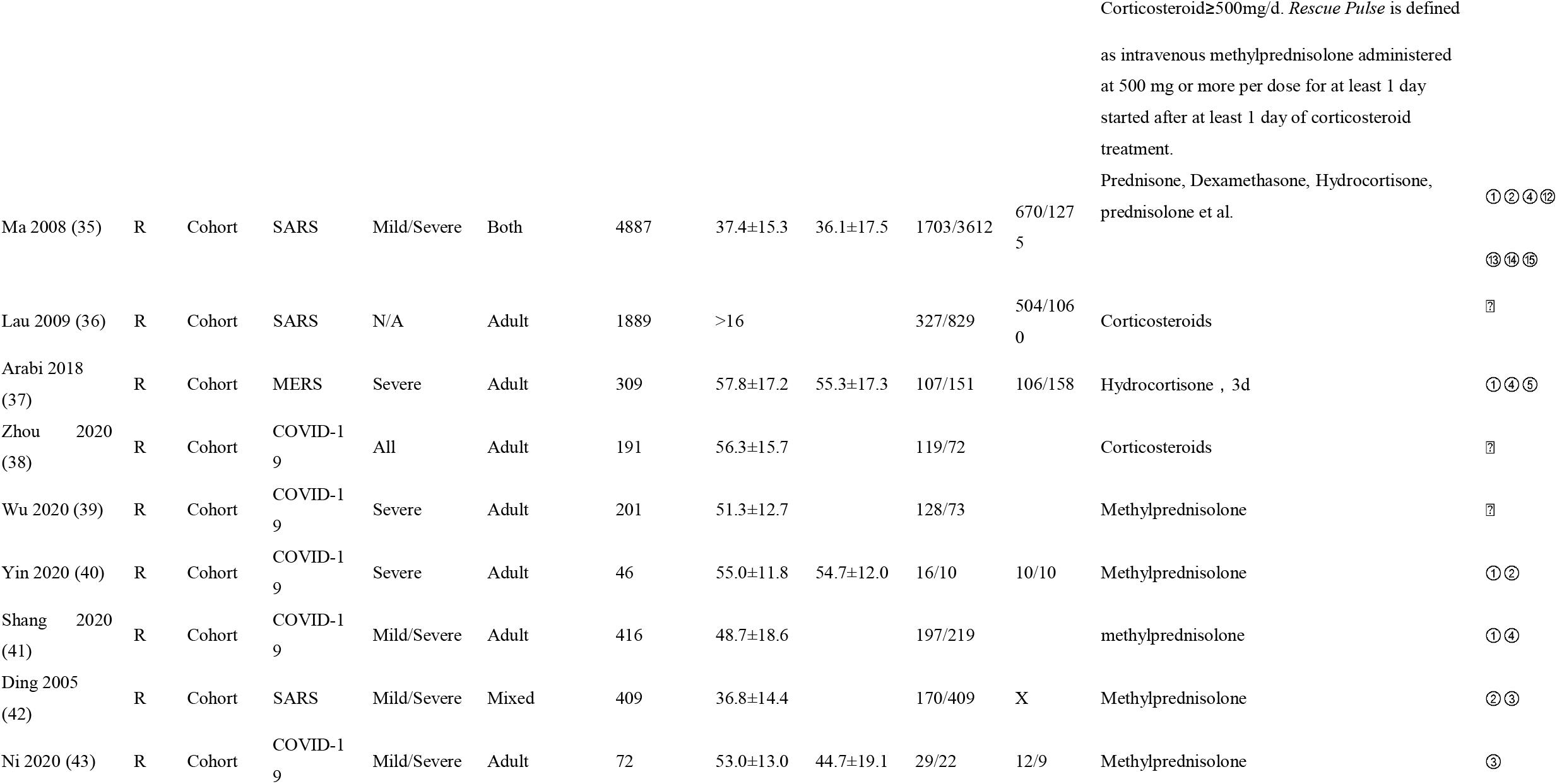
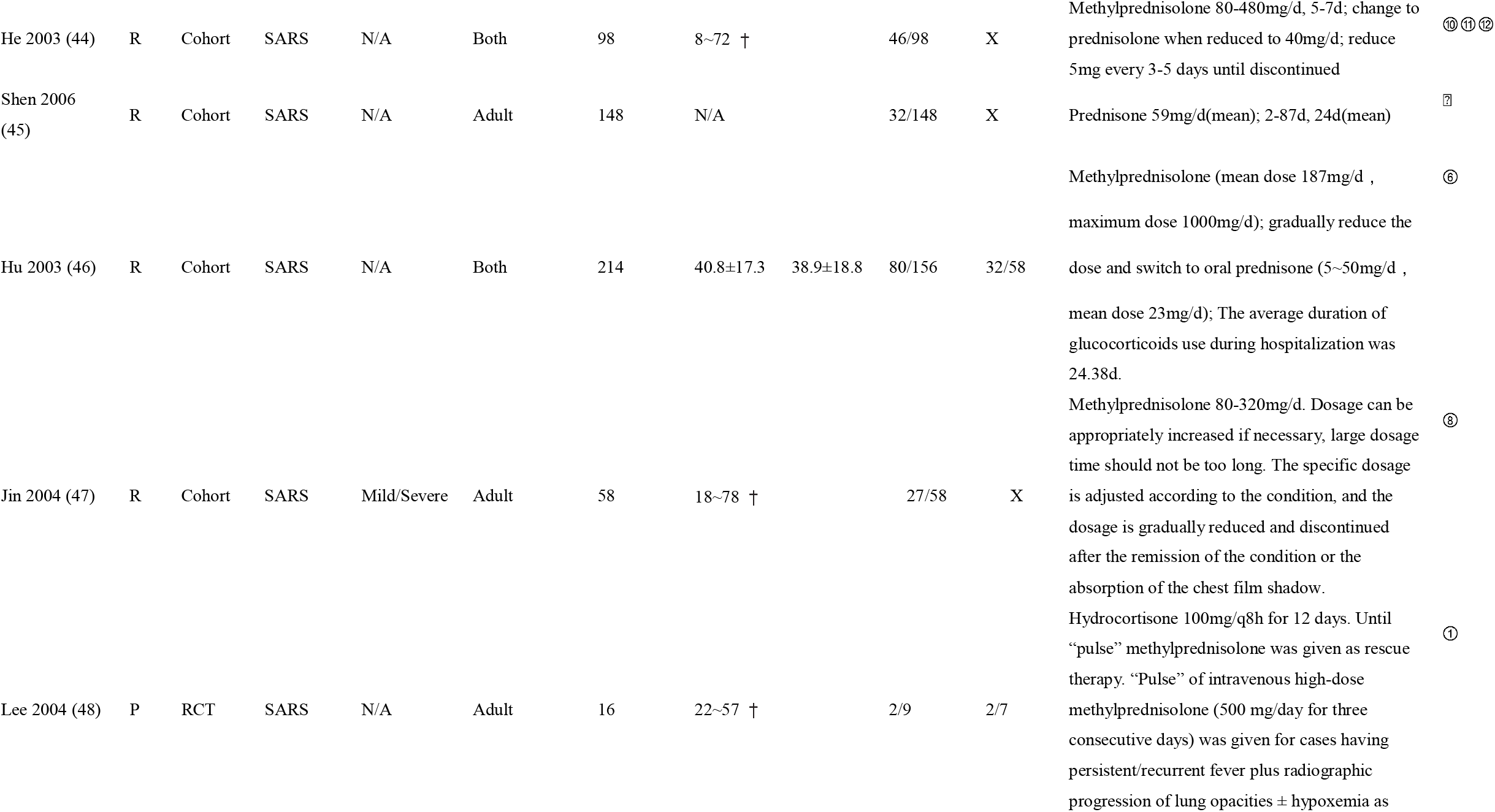
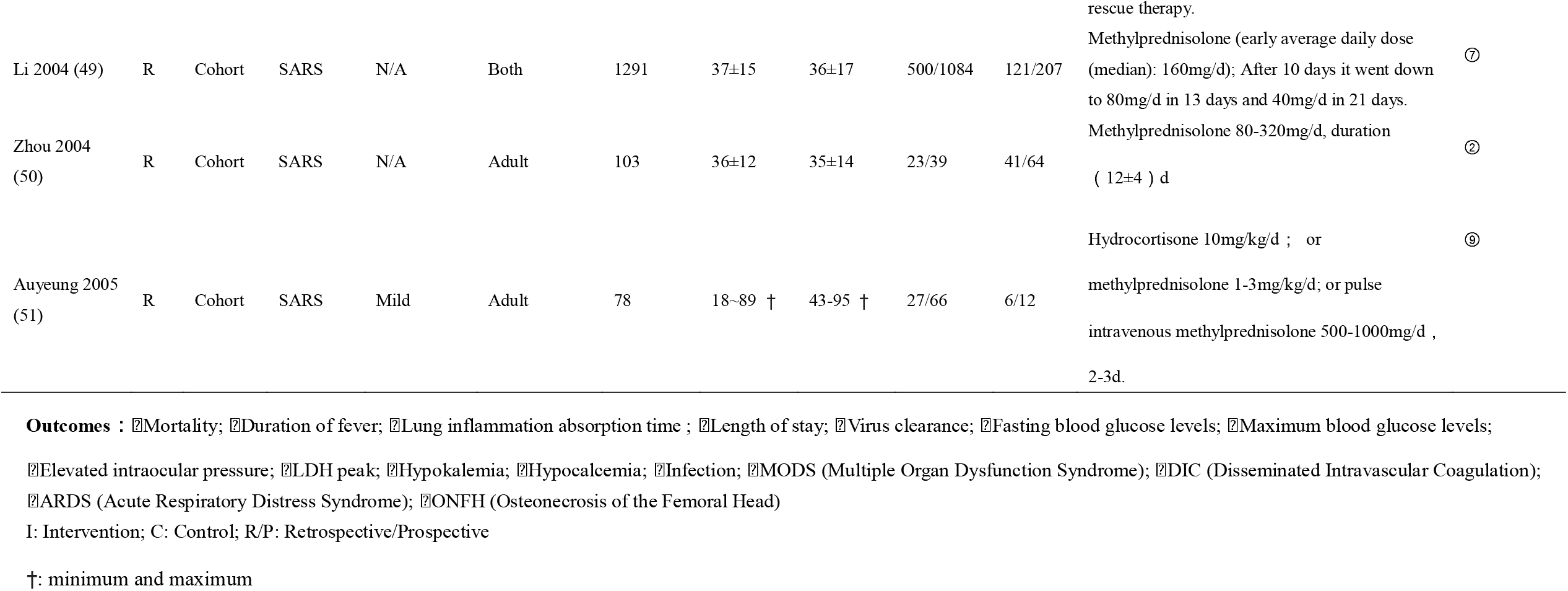
Characteristics of included studies

**Table 2.**
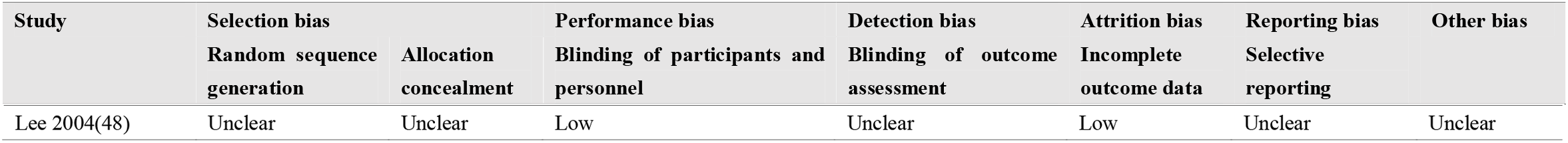
Assessment of risk of bias in RCT

**Table 3.**
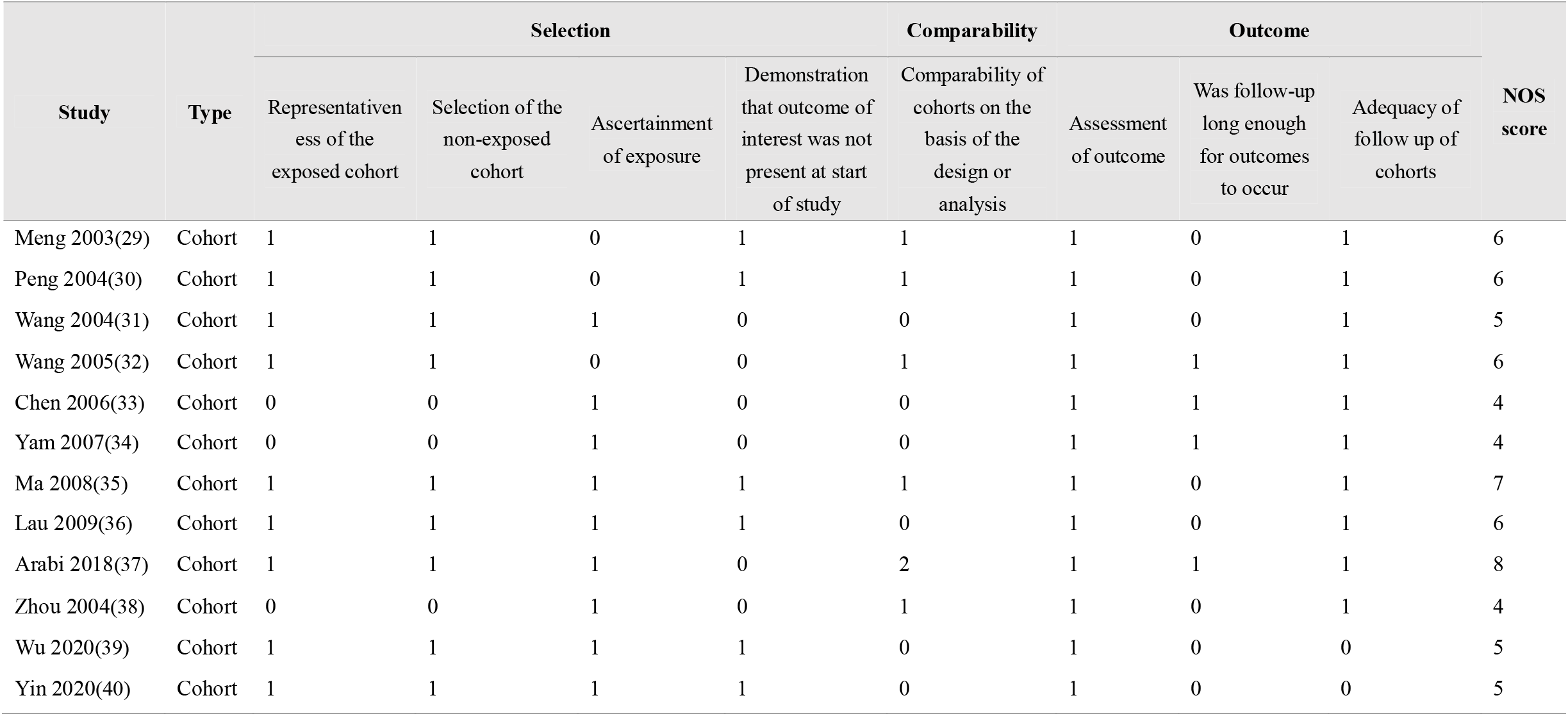
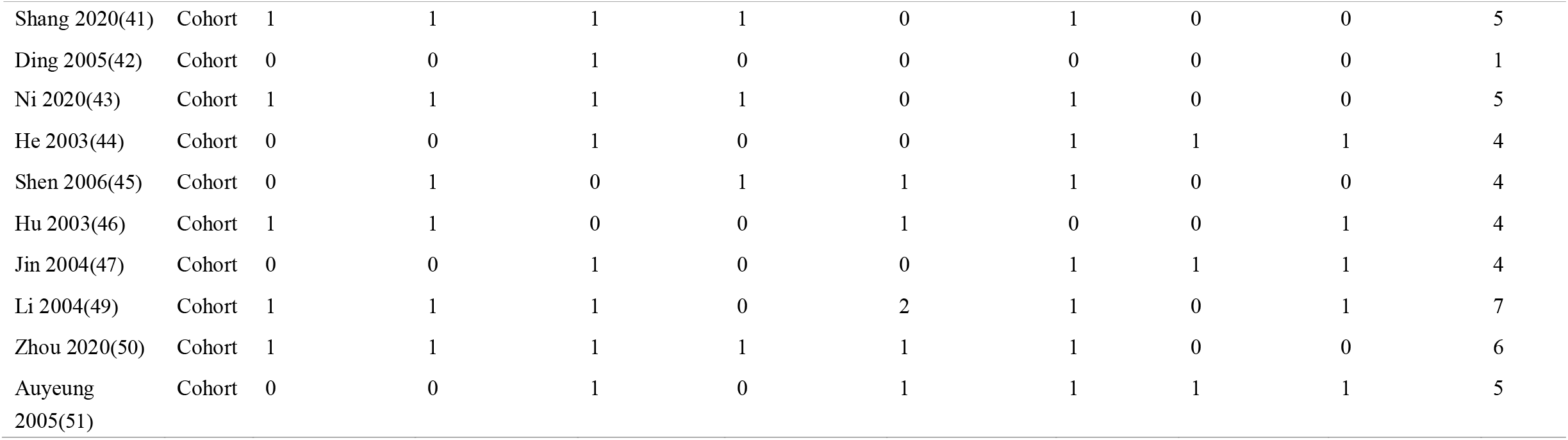
Assessment of risk bias in cohort studies

**Table 4.**
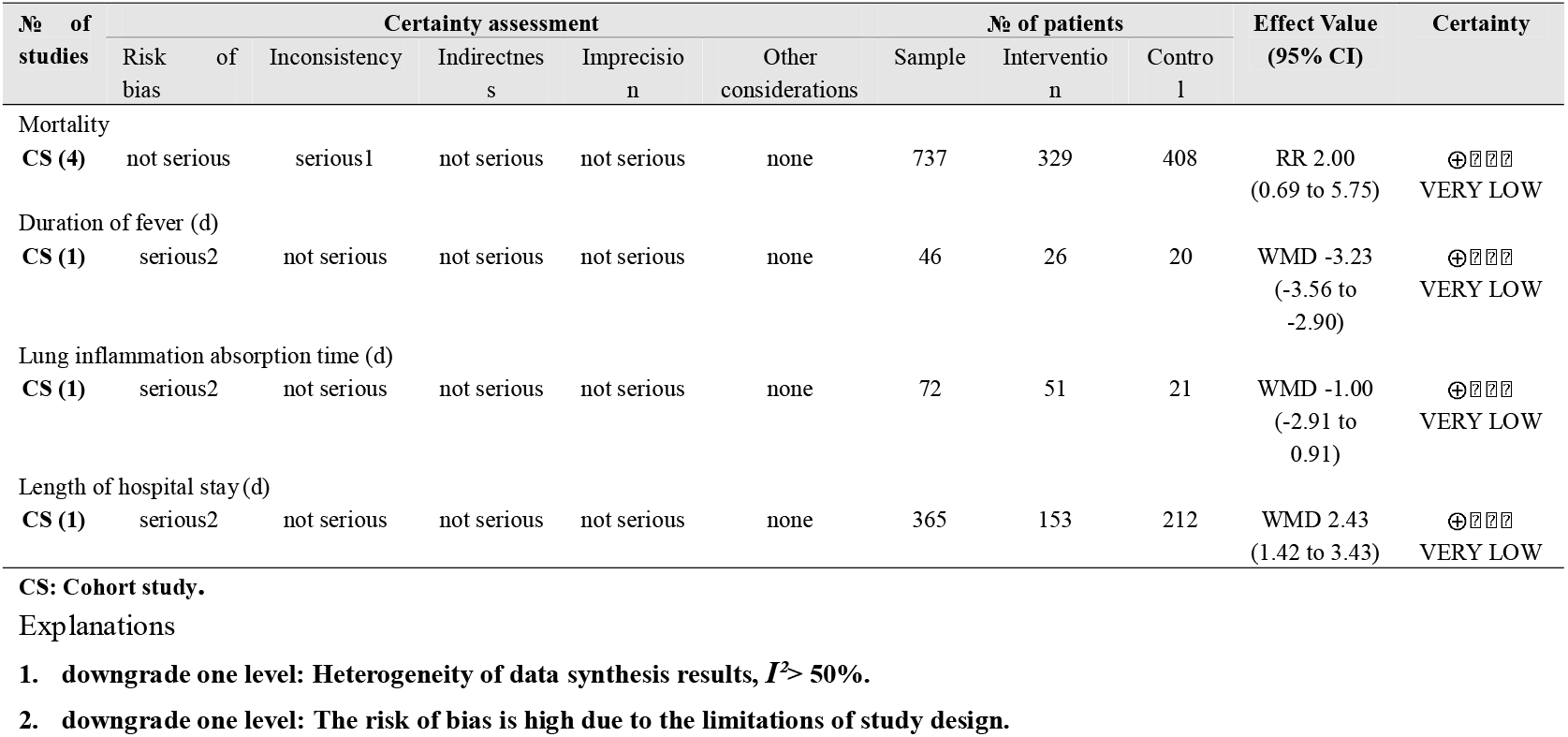
GRADE evidence profile of COVID

**Table 5.**
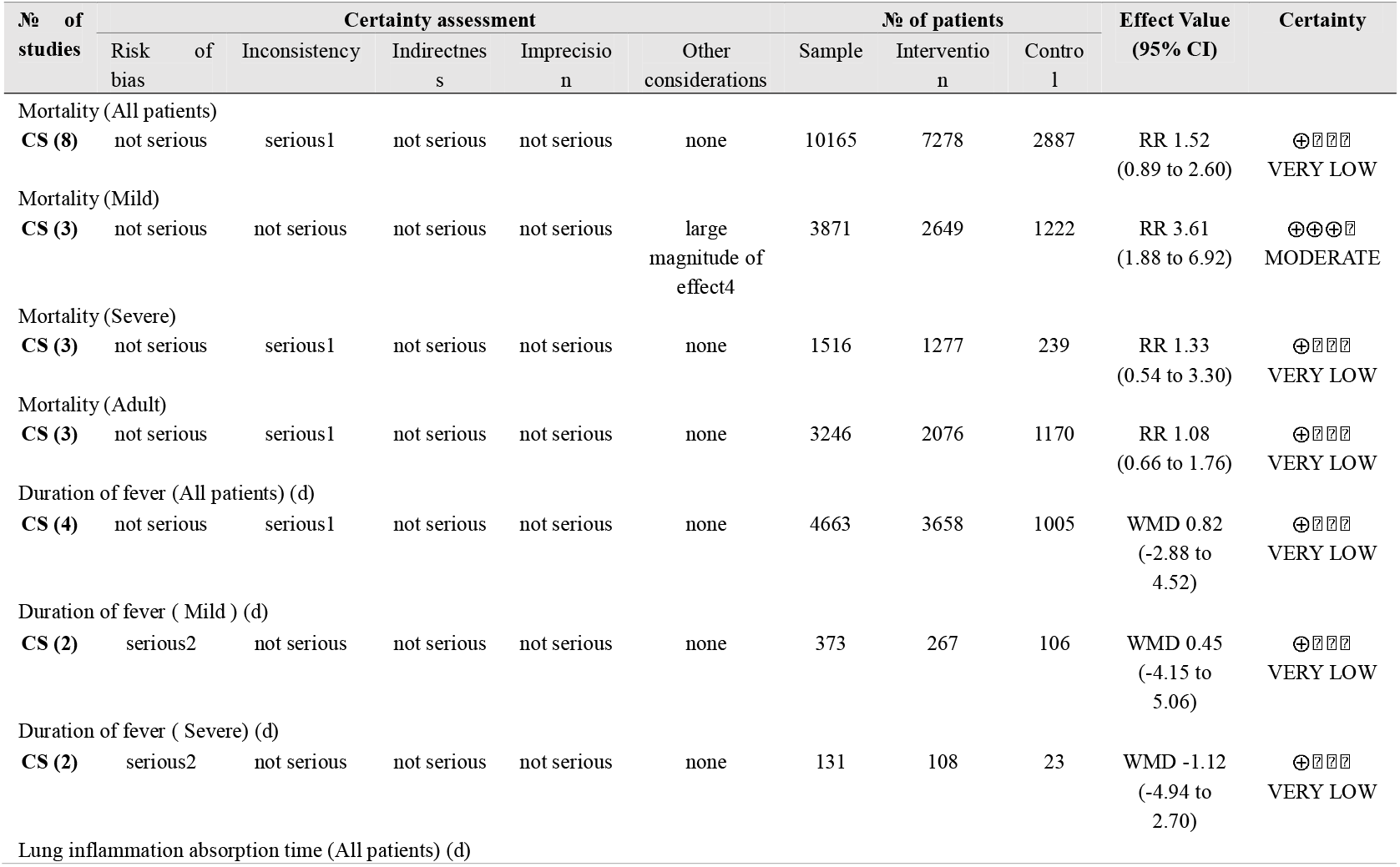
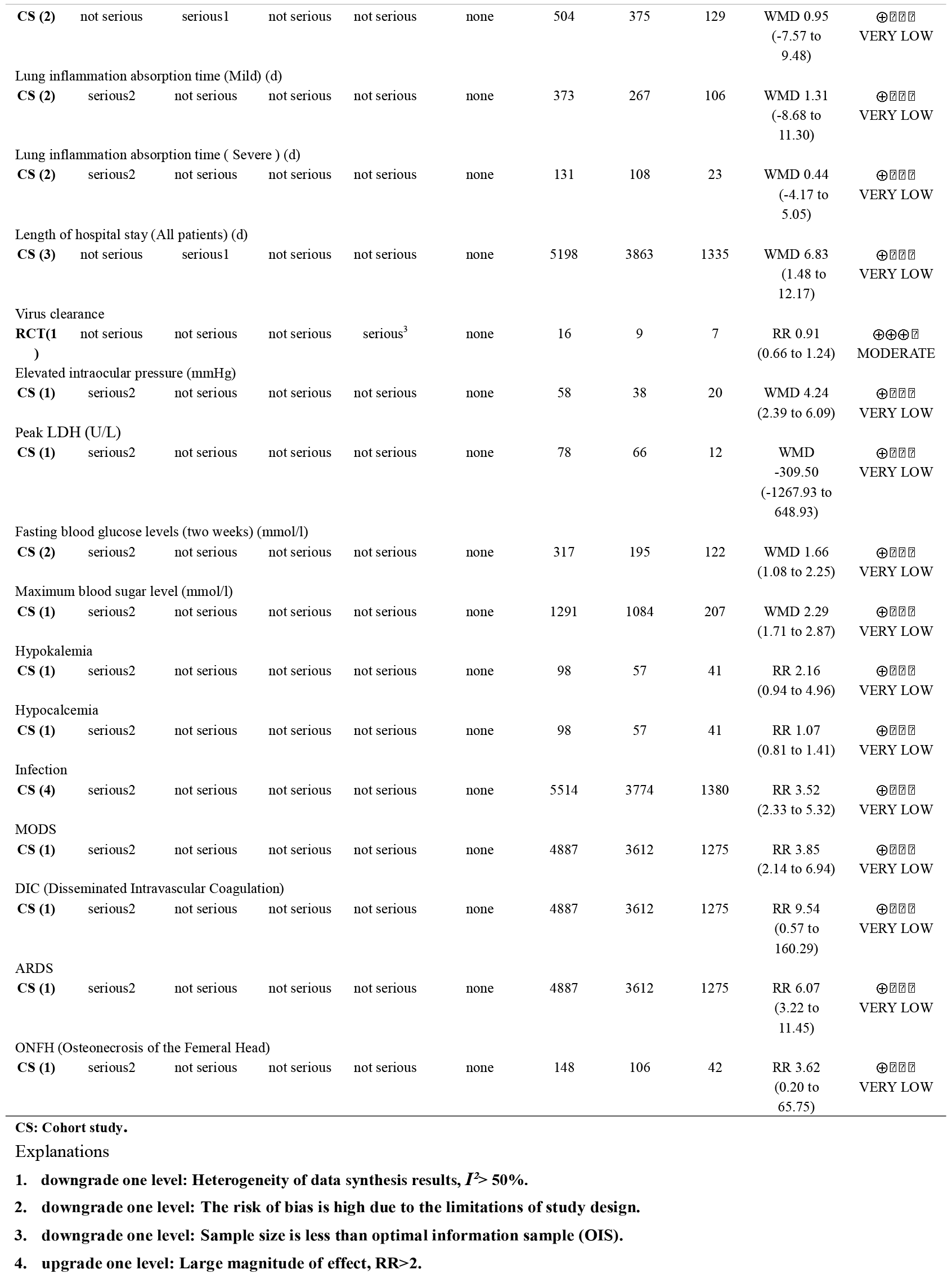
GRADE evidence profile of SARS studies

**Table 6.**
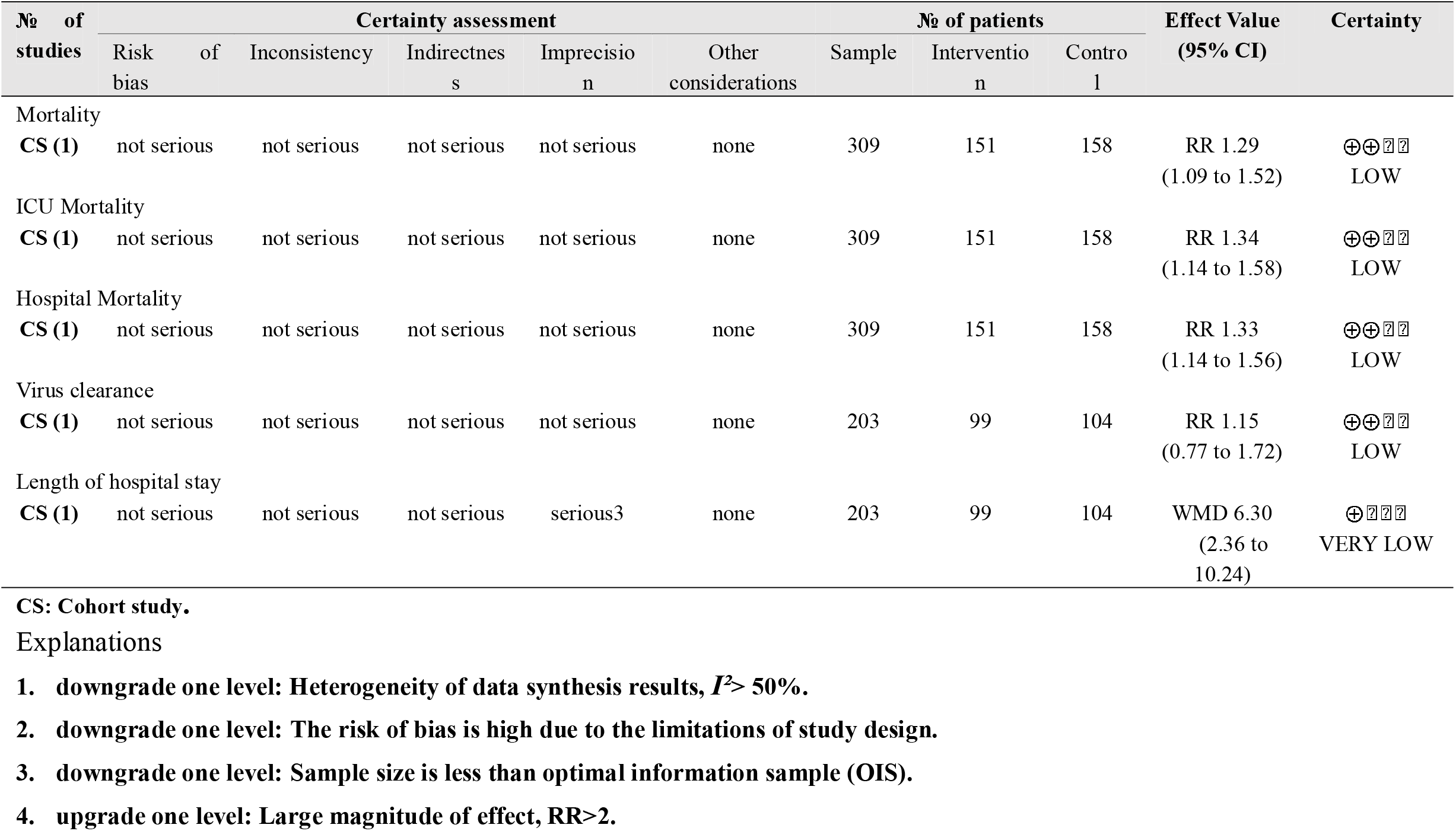
GRADE evidence profile of MERS studies

**Figure 1.**
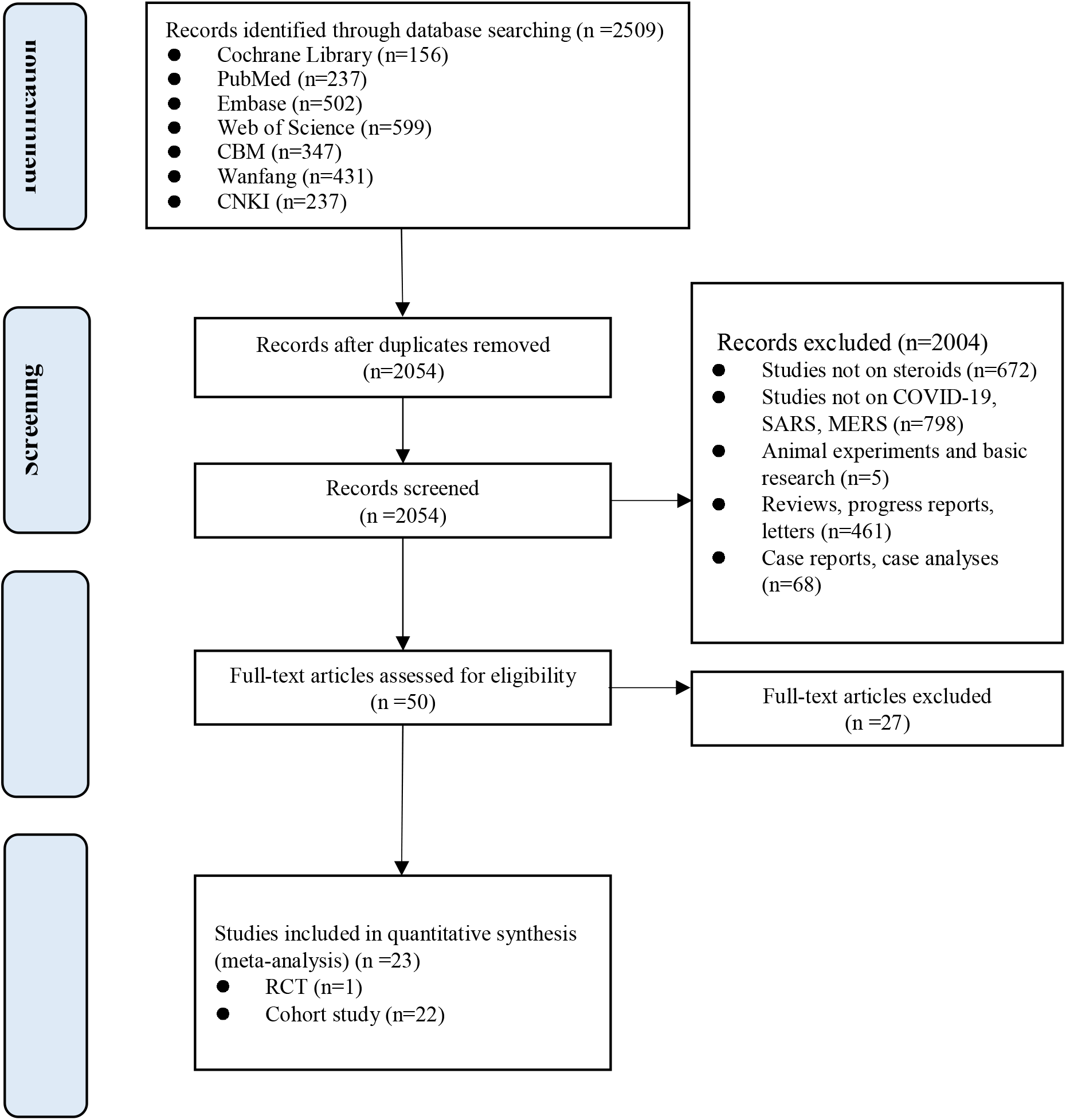
PRISMA flow chart.

### Meta-analyses

Mortality was assessed in thirteen cohort studies (four on COVID-19, eight on SARS, one on severe MERS) (29-41) with a total of 11,211 patients. The use of systemic glucocorticoid did not reduce the risk of death in COVID-19 (RR=2.0, 95% CI: 0.7 to 5.8, *I*^*2*^=90.9%) or SARS patients (RR=1.5, 95% CI: 0.9 to 2.6, *I*^*2*^=84.6%). Subgroup analyses showed that glucocorticoids also did not reduce the risk of death in severe cases of SARS (RR=1.3, 95% CI: 0.5 to 3.3, *I*^*2*^=67.4%) or in adults (RR=1.1, 95% CI: 0.7 to 1.8, *I*^*2*^=68.7%). In mild cases of SARS glucocorticoids even increased the risk of death (RR=3.6, 95% CI: 1.9 to 6.9). In adult patients with MERS, the use of glucocorticoid increased mortality (RR=1.3, 95% CI: 1.1 to 1.5) (*Figure 2, Figure 3, Figure 4*).

**Figure 2.**
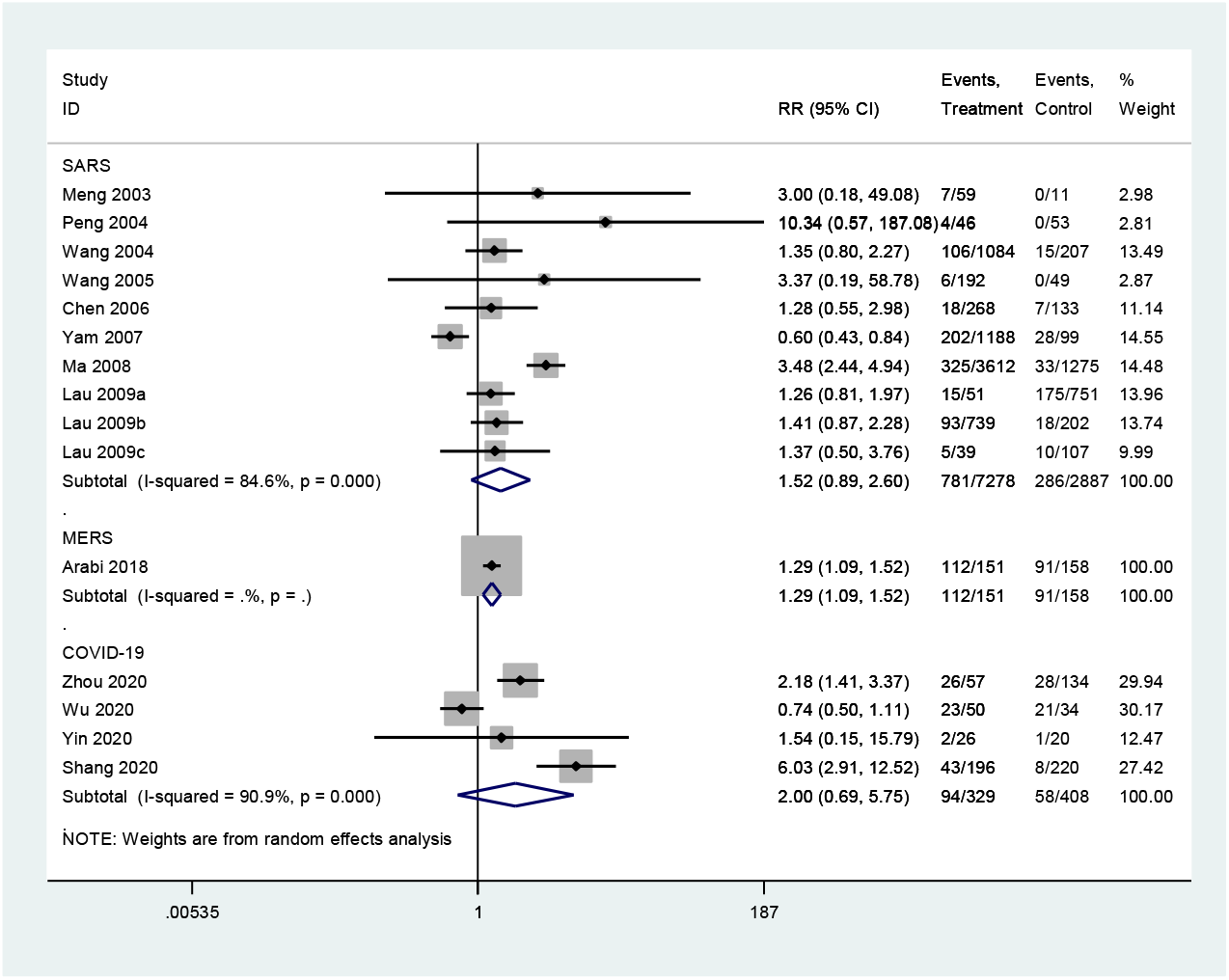
Relative risk of death in patients receiving versus not receiving glucocorticoid therapy: all patients.

**Figure 3.**
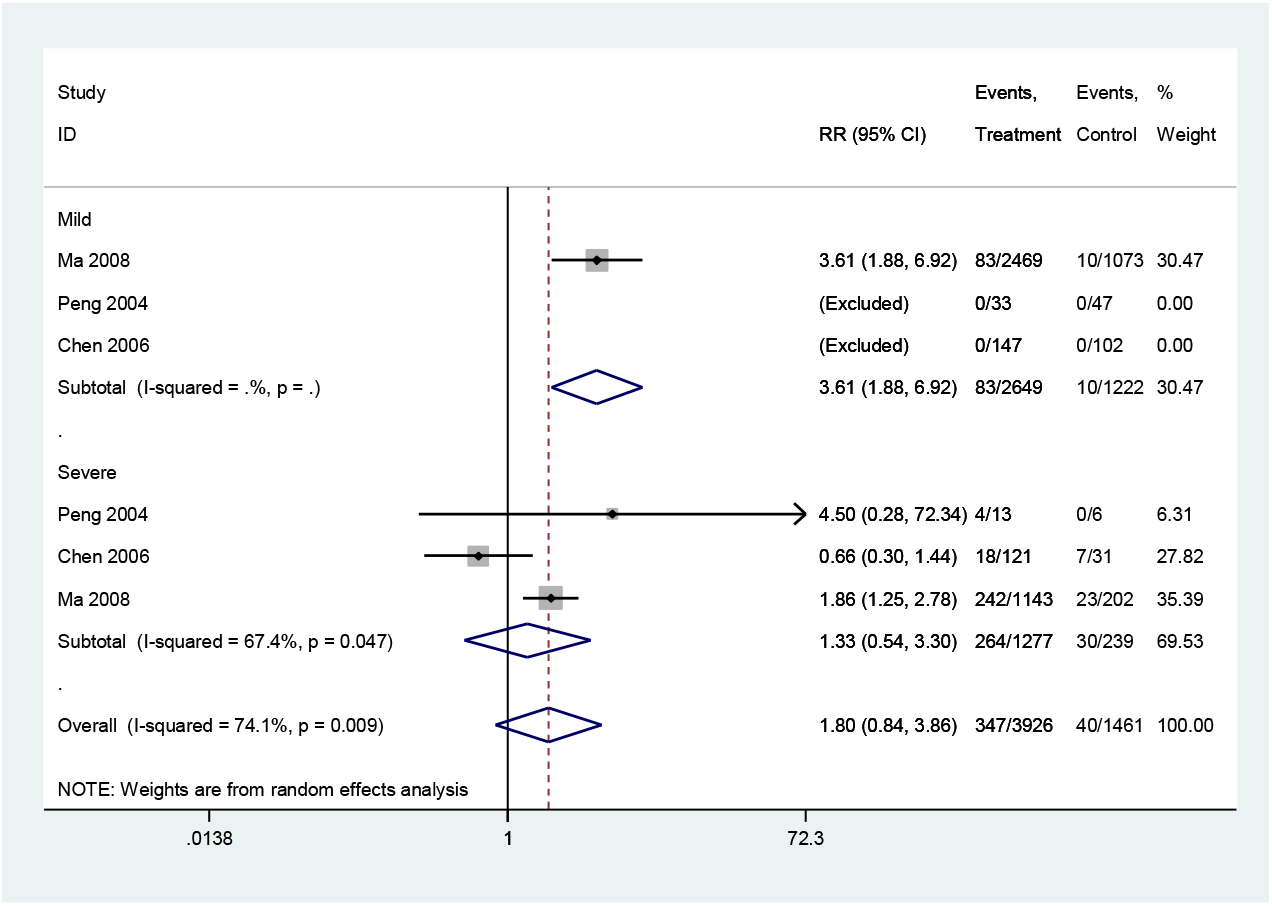
Relative risk of death in patients receiving versus not receiving glucocorticoid therapy: subgroup analyses of patients with mild and severe SARS.

**Figure 4.**
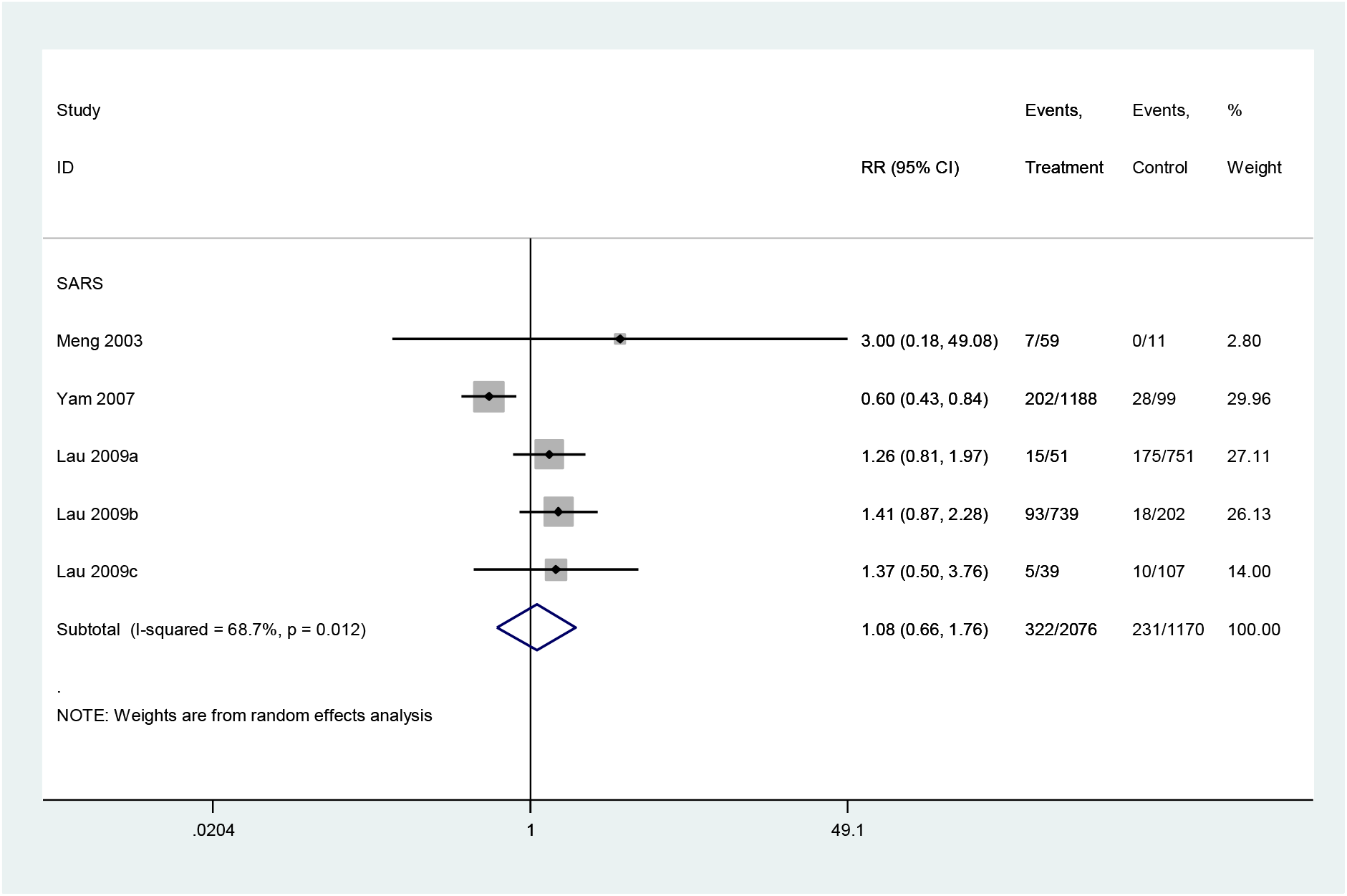
Relative risk of death in patients receiving versus not receiving glucocorticoid therapy: subgroup analyses of adult patients with SARS.

Five cohort studies with a total of 4709 patients assessed the duration of fever in COVID-19 (one study) and SARS (four studies) patients (30,32,35,40,42). The duration of fever was significantly lower in COVID-19 patients who received glucocorticoid treatment than in patients who received no glucocorticoid treatment (WMD=-3.2 d, 95% CI: −3.6 to −2.9), while for SARS patients there was no difference (WMD=0.8 d, 95% CI: −2.9 to 4.5, *I*^*2*^=97.9%). Subgroup analysis showed that glucocorticoid use did not shorten the duration of fever neither in patients with severe SARS (WMD=-1.1 d, 95% CI: −4.9 to 2.7, *I*^*2*^=58.3%) nor in patients with mild SARS (WMD=0.5 d, 95% CI: −4.2 to 5.1, *I*^*2*^=95.1%) (*Figure 5, Figure 6*).

**Figure 5.**
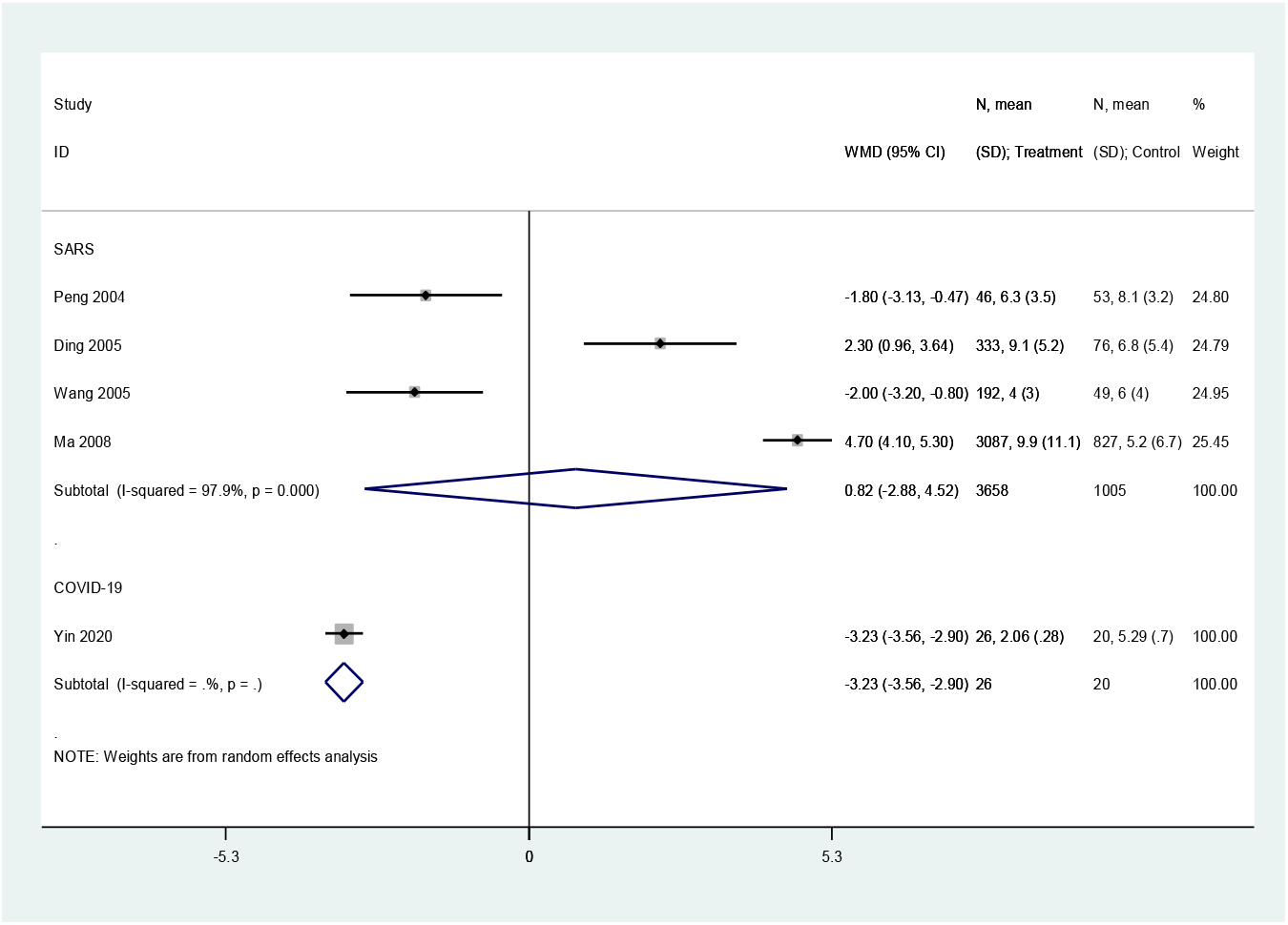
Duration of fever in patients receiving versus not receiving glucocorticoid therapy: all patients.

**Figure 6.**
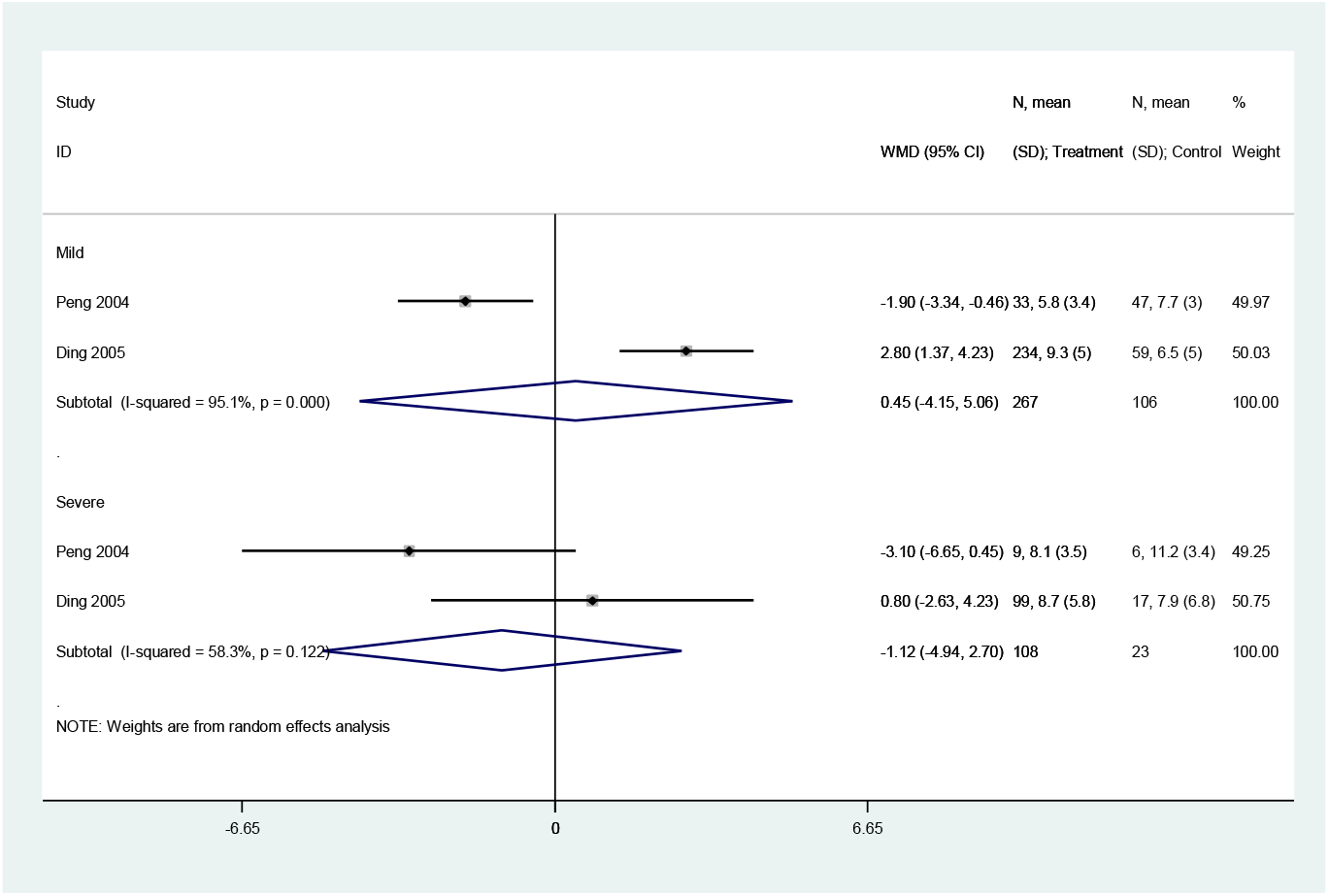
Duration of fever in patients receiving versus not receiving glucocorticoid therapy: subgroup analyses of patients with mild and severe SARS.

Three cohort studies assessed the duration of lung inflammation absorption in patients with COVID-19 (one study) and SARS (two studies) (30,42,43). No difference between patients who received or did not receive glucocorticoid treatment was found in neither COVID-19 (WMD=-1.0 d, 95% CI: −2.9 to 0.9) nor SARS (WMD=1.0 d, 95% CI: −7.6 to 9.5, *I*^*2*^=94.6%) patients. Subgroup analyses showed that using glucocorticoids did not shorten the absorption time of lung inflammation in patients with severe SARS regardless of severity (WMD=0.4 d, 95%CI: −4.2 to 5.1, *I*^*2*^=0%), or with mild SARS (WMD=1.3 d, 95%CI:-8.7 to 11.3, *I*^*2*^=95.5%) (*Figure 7, Figure 8*).

**Figure 7.**
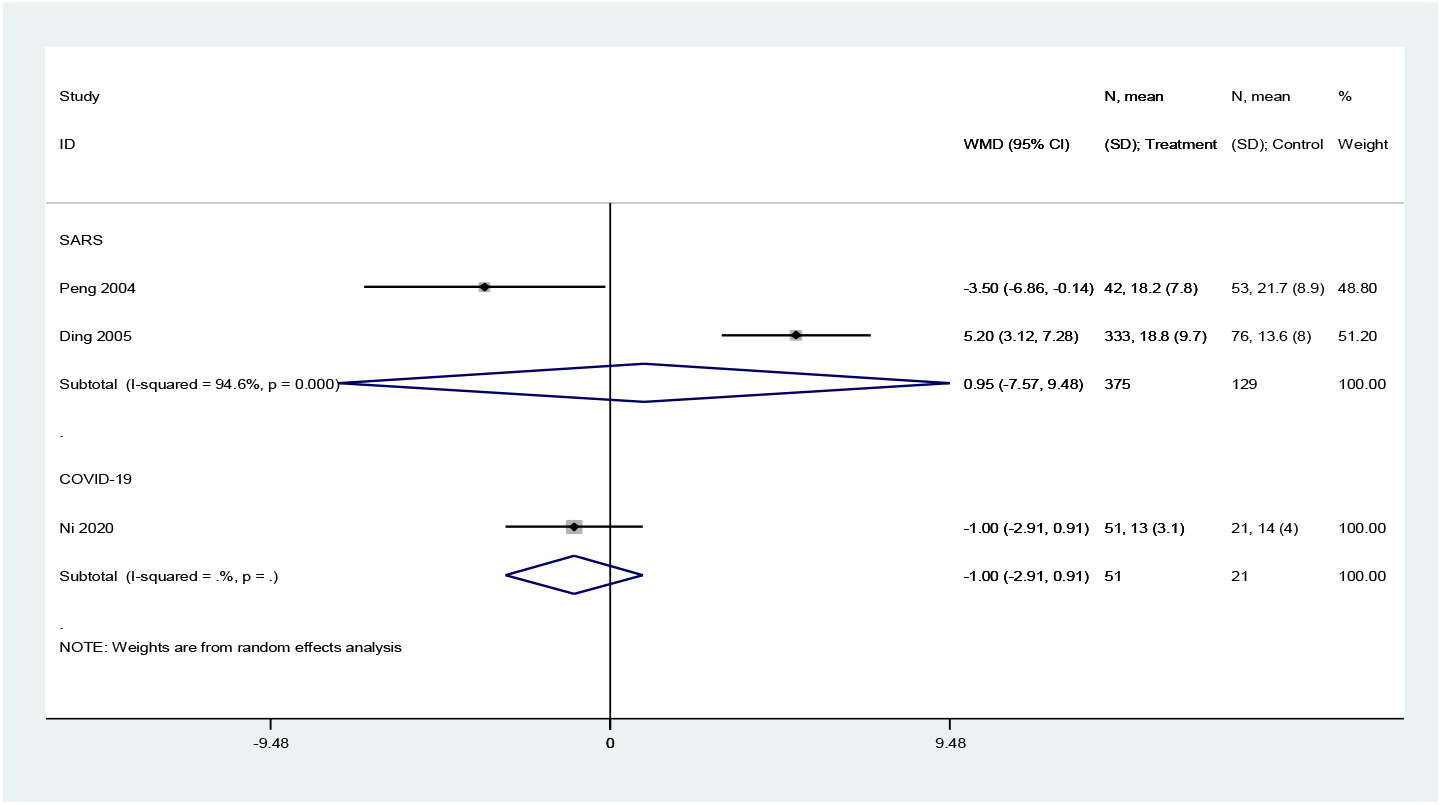
Lung inflammation absorption time in patients receiving versus not receiving glucocorticoid therapy: all patients.

**Figure 8.**
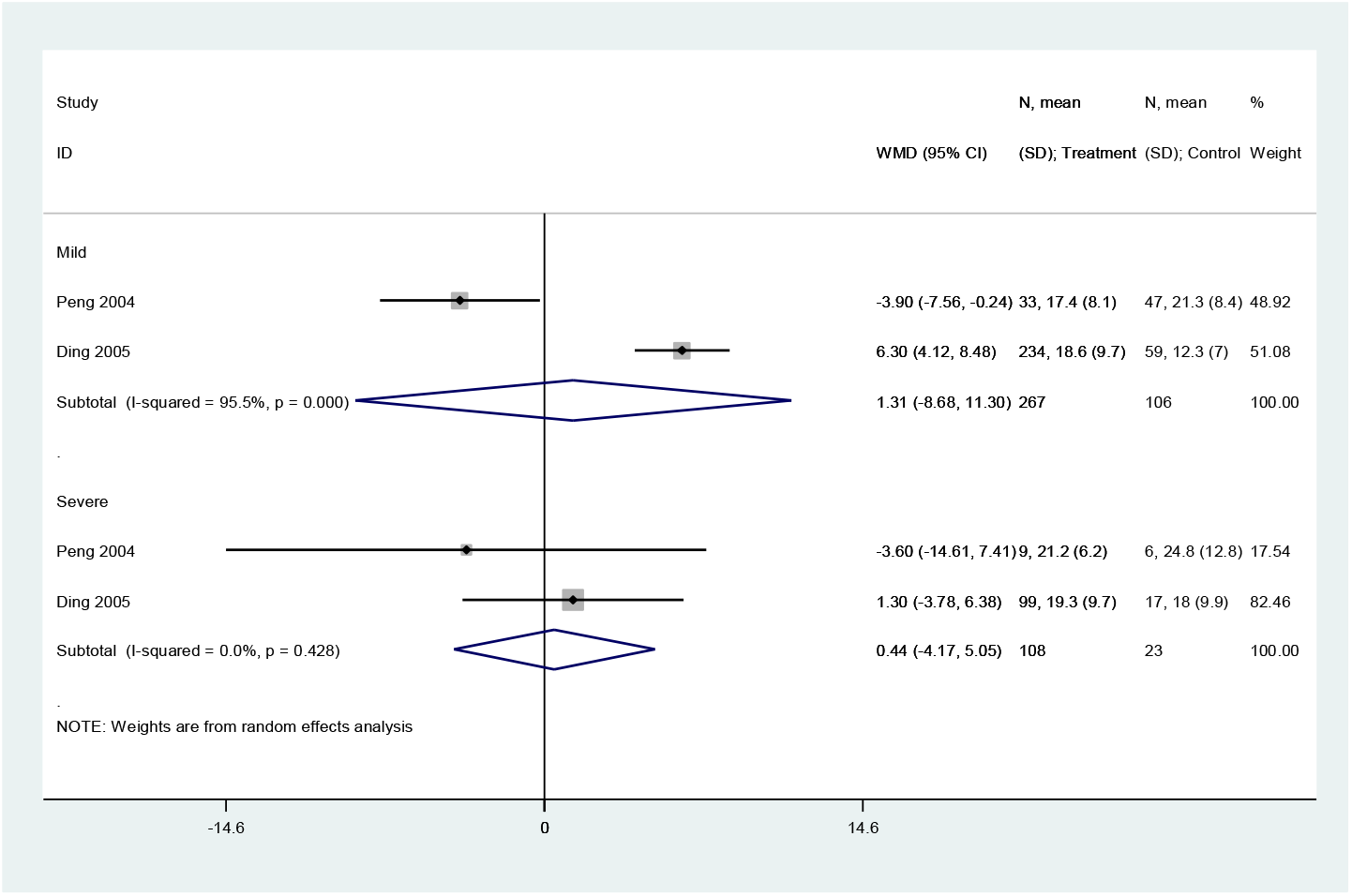
Lung inflammation absorption time in patients receiving versus not receiving glucocorticoid therapy: subgroup analyses of patients with mild and severe SARS.

Five cohort studies (one on COVID-19, three on SARS, one on severe MERS) with a total of 5872 patients assessed the duration of hospital stay (29,31,35,37,41). Patients treated with glucocorticoids stayed longer in the hospital than patients who did not receive glucocorticoids (COVID-19: WMD=2.4 d, 95%CI: 1.4 to 3.4, *I*^*2*^=0.0%; SARS: WMD=6.8 d, 95%CI: 1.5 to 12.2, *I*^*2*^=94.2%; MERS: WMD=6.3 d, 95%CI: 2.4 to 10.2) (*Figure 9*).

**Figure 9.**
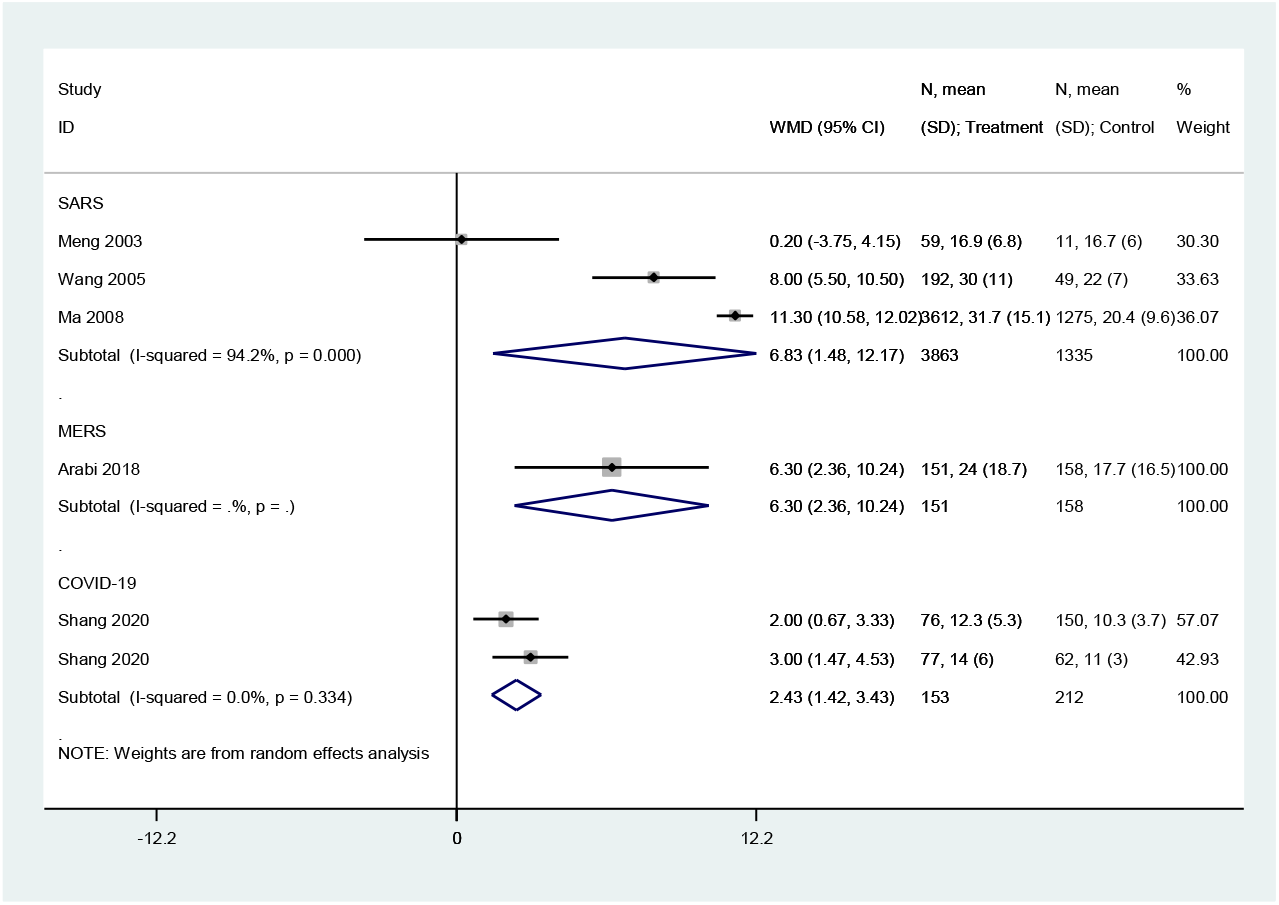
Length of stay in patients receiving versus not receiving glucocorticoid therapy: all patients.

Five cohort studies with a total of 5302 patients assessed the adverse outcomes in patients with SARS (29, 30, 35, 44, 45). Glucocorticoid use increased the risk of coinfections (bacterial or fungal) (RR=3.5, 95% CI: 2.3 to 5.3, *I*^*2*^=0%), MODS (RR=3.9, 95% CI: 2.1 to 6.9), and Acute Respiratory Distress Syndrome (ARDS) (RR=6.1, 95% CI: 3.2 to 11.5), while no significant association on DIC, hypokalemia, hypocalcemia and ONFH was found (*Figure 10*).

**Figure 10.**
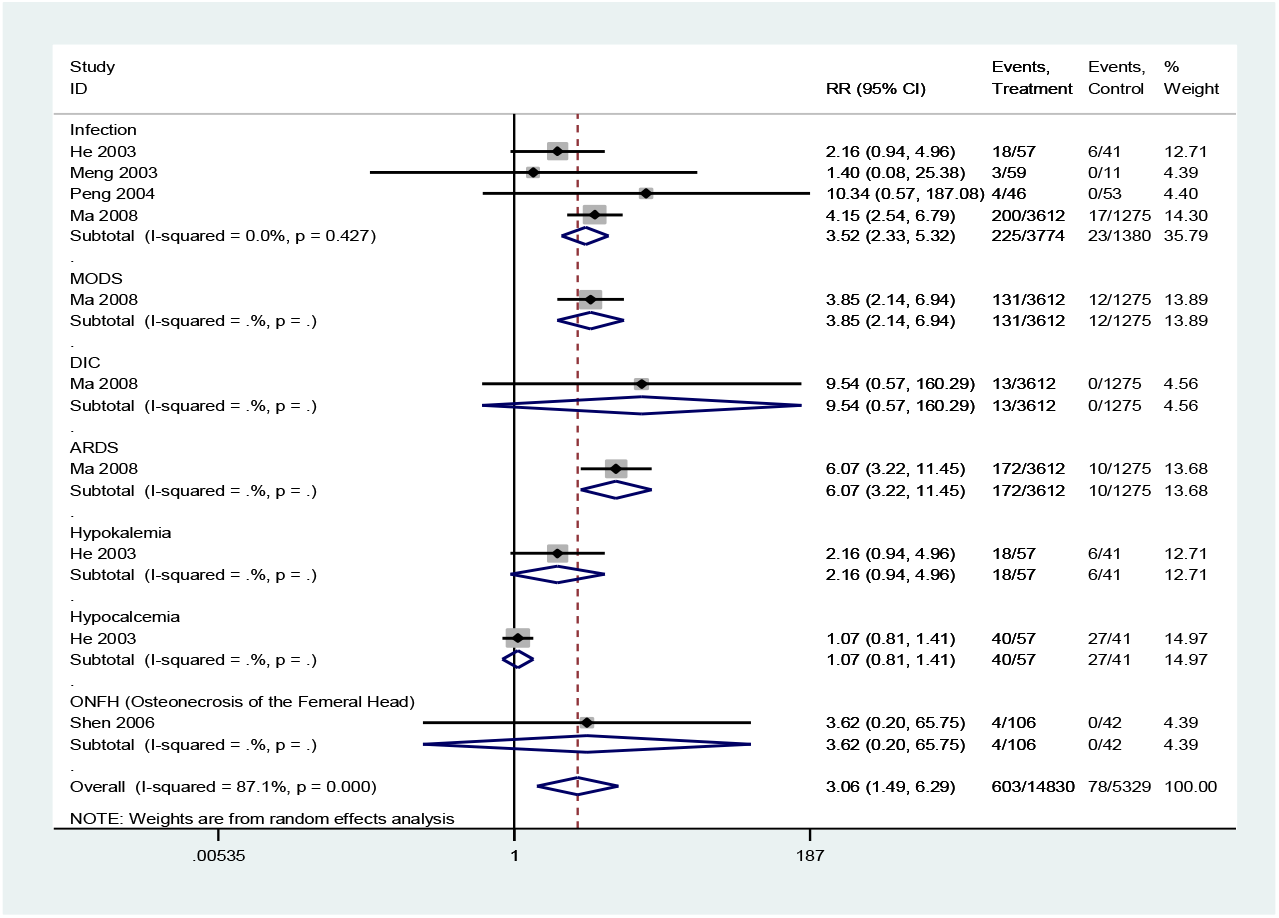
Relative risk of adverse events in patients receiving versus not receiving glucocorticoid therapy.

### Quality of evidence

The quality of evidence on the results on mortality in COVID-19 and SARS studies, of very low quality, and in MERS studies of low quality (*Tables 4-6*).

### Sensitivity Analysis

Heterogeneity among studies on SARS assessing mortality was significant (*I*^*2*^*=*84.6%). Heterogeneity in studies of mortality in SARS patients was reduced to 67.4% in the subgroup analysis of severe cases, and to 68.7% in the subgroup analysis of adults. Therefore, disease severity and age are probably the main sources of heterogeneity in the meta-analysis of mortality. We conducted a sensitivity analysis on the on the SARS mortality by omitting one study at a time. Two studies had a significant impact on the results of the meta-analysis (34, 35) (*Figure 11*). The dosing of glucocorticoids was different in the study by Yam *et al* than in other studies, so the high heterogeneity in the meta-analysis on mortality may be at least partly caused by the different dosing (34).

**Figure 11.**
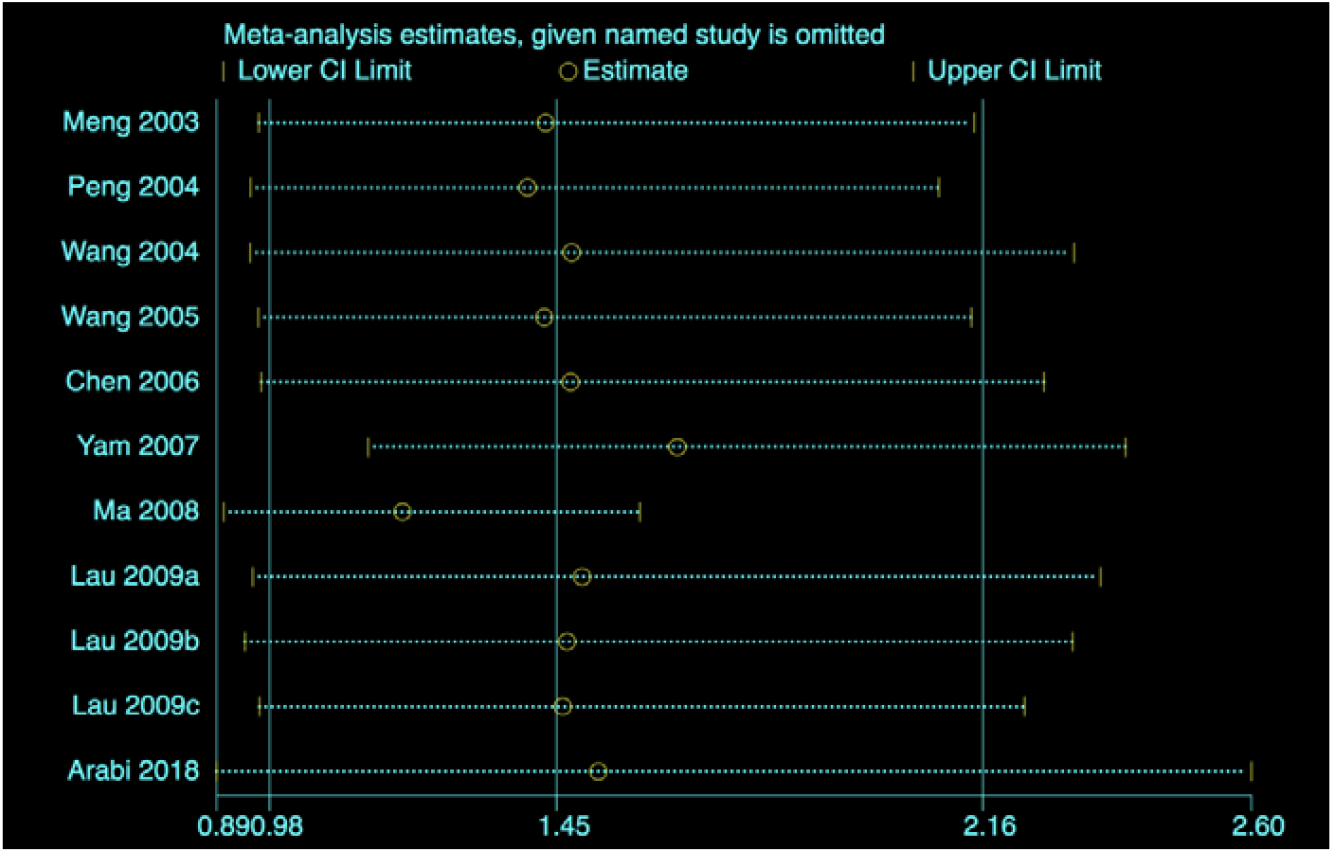
Sensitivity analysis of mortality in SARS patients.

### Publication Bias

We assessed publication bias was for the eight studies on SARS mortality. The Egger regression test showed that publication bias was unlikely (*P*=0.619) (***Figure 12***).

**Figure 12.**
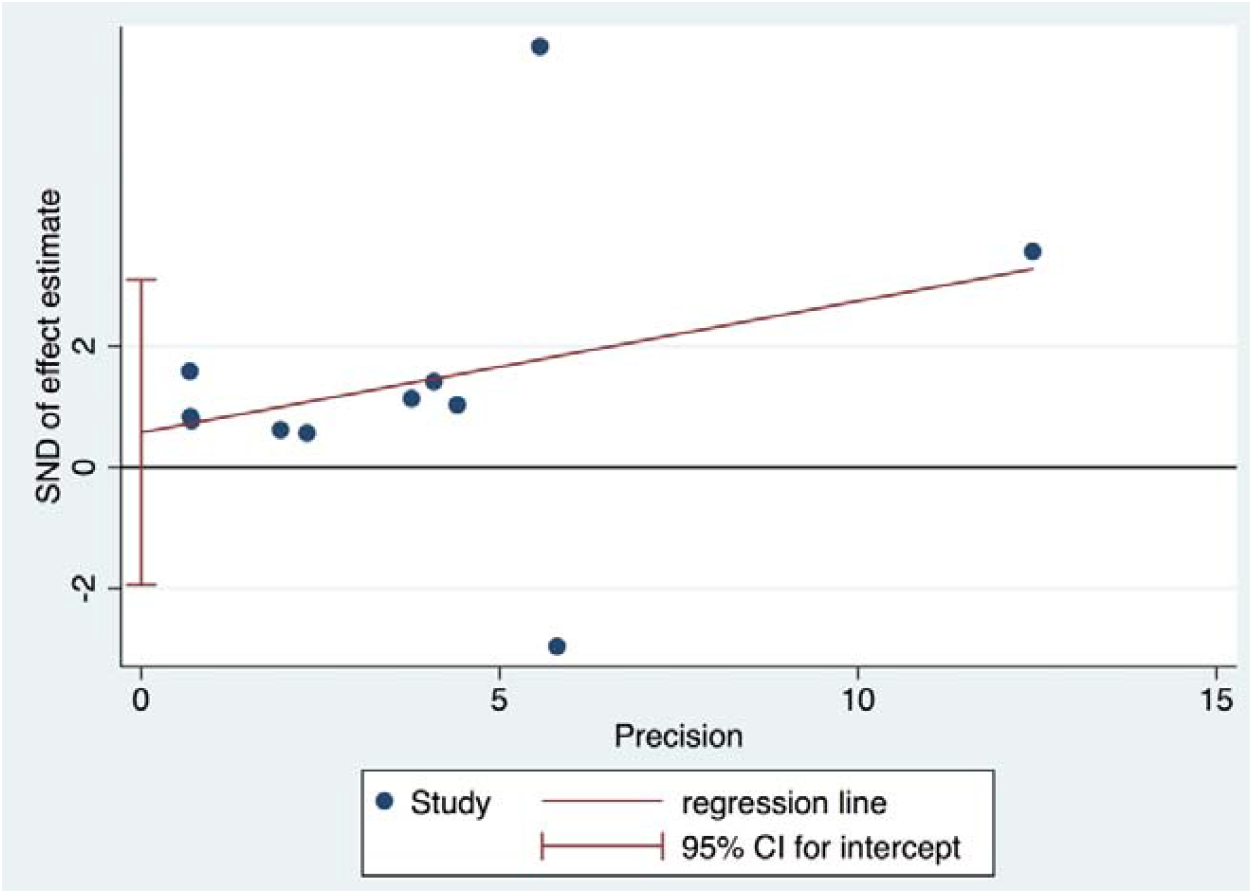
Publication bias (Egger test)

## Discussion

Our study identified direct evidence on the clinical efficacy of glucocorticoid therapy for five difference outcomes in adults with COVID-19. Evidence of low to very low-quality showed that glucocorticoid therapy significantly reduced the duration of fever, but not the risk of death and lung inflammation absorption in patients with COVID-19 or SARS. In addition, glucocorticoid therapy may even prolong the duration of hospitalization. Long-term use of high-dose glucocorticoids increased the risk of adverse reactions such as infections and osteonecrosis. We found moderate-quality evidence that in patients with mild SARS glucocorticoids may be associated with a more than three-fold increase in the risk of death.

Systemic glucocorticoids are highly effective anti-inflammatory drugs, but their use against SARS-CoV-2 infection remains controversial. A case series of children with COVID-19 in Hubei reported that systematic glucocorticoids (dose 2 mg/kg) were given to both two included critical cases in combination with invasive mechanical ventilation and intravenous immunoglobulin. In both children, the symptoms on admission were alleviated, although in one of them only partly (52). A recent cohort study from JAMA Internal Medicine reported that among COVID-19 patients with ARDS, treatment with methylprednisolone decreased the risk of death (39). Our results are compared with published systematic reviews of glucocorticoid therapy for severe pneumonia. A recent rapid review of COVID-19 treatment showed controversial evidence on the use of corticosteroids, and could not give any suggestion on the use of corticosteroids due to the lack of quantitative synthesis (53). A systematic review covering in vitro studies on SARS, SARS in humans and other diseases such as ARDS, found that 25 of the 29 included studies were inconclusive, and the remaining four found glucocorticoids harmful (54). A recent systematic review of influenza pneumonia showed that glucocorticoid therapy increased the risk of death (RR=1.75, 95% CI: 1.30-2.36), length of ICU stay (RR=2.14 days, 95% CI: 1.17-3.10), and risk of secondary infections (RR=1.98, 95% CI: 1.04-3.78) (55). A meta-analysis of SARS in 2017 showed that the incidence of osteonecrosis increased with the dosing of systemic glucocorticoids, and the summary RR of osteonecrosis was 1.57 (95% CI 1.30–1.89) (56). As retrospective studies have shown that the glucocorticoids were given for 19% to 26% of patients with COVID-19 (and to 45% of patients with severe disease), there is a high risk that this therapy is currently misused (57-59). In summary, the current research evidence does not support the routine use of systemic glucocorticoids for patients with COVID-19. Because COVID-19 tends to be less severe in children than in adults (60), the use of systemic glucocorticoids should in particular not be recommended in children.

### Strengths and Limitations

This is to our knowledge the first comprehensive and systematic review of the effectiveness and safety of glucocorticoid therapy for patients with COVID-19 using data from the COVID-19 studies, and can be considered at the moment as the best evidence for decision-making on this topic. We conducted meta-analyses to quantitatively synthesize the findings of the included studies and objectively evaluate the current research evidence. Our study had also several limitations. We found only limited direct evidence of systemic glucocorticoids therapy for COVID-19, and had to use indirect evidence from the SARS and MERS epidemics. We also could not conduct subgroup analyses according to the dose and type of glucocorticoids because of the small number of studies.

## Conclusions

In conclusion, glucocorticoid therapy may increase the risk of death in patients with coronavirus infections who have mild symptoms. We found no association between glucocorticoids and mortality in patients with severe symptoms. In the context of clinical trials, low dose systemic glucocorticoid therapy for a short duration may be acceptable.

## Data Availability

The data used to support the findings of this study are included within the article.

## Author contributions

(I) Conception and design: Lu S, Zhou Q and Huang L; (II) Administrative support: Y Chen; (III) Provision of study materials or patients: Lu S, Huang L, Zhou Q; (IV) Collection and assembly of data: Lu S, Zhou Q and Shi Q; (V) Data analysis and interpretation: Zhou Q, Lu S and Zhao S; (VI) Manuscript writing: All authors; (VII) Final approval of manuscript: All authors.

## Funding

This work was supported by grants from National Clinical Research Center for Child Health and Disorders (Children’s Hospital of Chongqing Medical University, Chongqing, China) (grant number NCRCCHD-2020-EP-01); Special Fund for Key Research and Development Projects in Gansu Province in 2020; The fourth batch of “Special Project of Science and Technology for Emergency Response to COVID-19” of Chongqing Science and Technology Bureau. Special funding for prevention and control of emergency of COVID-19 from Key Laboratory of Evidence Based Medicine and Knowledge Translation of Gansu Province (grant number No. GSEBMKT-2020YJ01).

## Acknowledgments

We thank Janne Estill, Institute of Global Health of University of Geneva for providing guidance and comments for our review. We thank all the authors for their wonderful collaboration.

## Footnote

### Conflicts of Interest

The authors have no conflicts of interest to declare.

### Ethical Statement

The authors are accountable for all aspects of the work in ensuring that questions related to the accuracy or integrity of any part of the work are appropriately investigated and resolved.

## Supplementary material

### Supplementary Material 1-Search strategy

#### PubMed

#1. “COVID-19”[Supplementary Concept]

#2. “Severe Acute Respiratory Syndrome Coronavirus 2”[Supplementary Concept]

#3. “Middle East Respiratory Syndrome Coronavirus”[Mesh]

#4. “Severe Acute Respiratory Syndrome”[Mesh]

#5. “SARS Virus”[Mesh]

#6. “COVID-19”[Title/Abstract]

#7. “SARS-COV-2”[Title/Abstract]

#8. “Novel coronavirus” [Title/Abstract]

#9. “2019-novel coronavirus” [Title/Abstract]

#10. “coronavirus disease-19” [Title/Abstract]

#11. “coronavirus disease 2019” [Title/Abstract]

#12. “COVID 19” [Title/Abstract]

#13. “Novel CoV” [Title/Abstract]

#14. “2019-nCoV” [Title/Abstract]

#15. “2019-CoV” [Title/Abstract]

#16. “Wuhan-Cov” [Title/Abstract]

#17. “Wuhan Coronavirus” [Title/Abstract]

#18. “Wuhan seafood market pneumonia virus” [Title/Abstract]

#19. “Middle East Respiratory Syndrome” [Title/Abstract]

#20. “MERS” [Title/Abstract]

#21. “MERS-CoV” [Title/Abstract]

#22. “Severe Acute Respiratory Syndrome” [Title/Abstract]

#23. “SARS” [Title/Abstract]

#24. “SARS-CoV” [Title/Abstract]

#25. “SARS-Related” [Title/Abstract]

#26. “SARS-Associated” [Title/Abstract]

#27. #1-#26/ OR

#28. “adrenal cortex hormones”[Mesh]

#29. “Beclomethasone” [Title/Abstract]

#30.” betamethasone valerate “[Mesh]

#31. “Budesonid”[Title/Abstract]

#32. “Cortodoxone”[Title/Abstract]

#33. “Dexamethasone”[Title/Abstract]

#34.” glucocorticoids”[Mesh]

#35. “Hydrocortisone”[Title/Abstract]

#36. “Hydroxycorticosteroids”[Title/Abstract]

#37.” methylprednisolone”[Mesh]

#38. “adrenal cortex hormone*”[Title/Abstract]

#39. “becl?met*”[Title/Abstract]

#40. “betamet?asone*”[Title/Abstract]

#41. “budesonide*”[Title/Abstract]

#42. “clobetasol* “[Title/Abstract]

#43. “corticoid*”[Title/Abstract]

#44. “corticosteroid*”[Title/Abstract]

#45. “corticosterone* “[Title/Abstract]

#46. “cortisone*”[Title/Abstract]

#47. “cortodoxone*”[Title/Abstract]

#48. “dexamet?asone*”[Title/Abstract]

#49. “glucocortico*”[Title/Abstract]

#50. “hydrocortisone*”[Title/Abstract]

#51. “hydroxycorticosteroid*”[Title/Abstract]

#52. “hydroxypregnenolone* “[Title/Abstract]

#53. “methylprednisolone*”[Title/Abstract]

#54. “prednisolone*”[Title/Abstract]

#55. “prednisone*”[Title/Abstract]

#56. “pregnenedione*”[Title/Abstract]

#57. “pregnenolone*”[Title/Abstract]

#58. “tetrahydrocortisol*”[Title/Abstract]

#59. “triamcinolone*”[Title/Abstract]

#60. #28-#59/ OR

#61. #27 AND#60

#### EMBASE

#1. ‘middle east respiratory syndrome coronavirus’/exp

#2. ‘severe acute respiratory syndrome’/exp

#3. ‘sars coronavirus’/exp

#4. ‘COVID-19’:ab,ti

#5. ‘SARS-COV-2’:ab,ti

#6. ‘novel coronavirus’:ab,ti

#7. ‘2019-novel coronavirus’:ab,ti

#8. ‘coronavirus disease-19’:ab,ti

#9. ‘coronavirus disease 2019’:ab,ti

#10. ‘COVID 19’:ab,ti

#11. ‘novel cov’:ab,ti

#12. ‘2019-ncov’:ab,ti

#13. ‘2019-cov’:ab,ti

#14. ‘wuhan-cov’:ab,ti

#15. ‘wuhan coronavirus’:ab,ti

#16. ‘wuhan seafood market pneumonia virus’:ab,ti

#17. ‘middle east respiratory syndrome’:ab,ti

#18. ‘middle east respiratory syndrome coronavirus’:ab,ti

#19. ‘mers’:ab,ti

#20. ‘mers-cov’:ab,ti

#21. ‘severe acute respiratory syndrome’:ab,ti

#22. ‘sars’:ab,ti

#23. ‘sars-cov’:ab,ti

#24. ‘sars-related’:ab,ti

#25. ‘sars-associated’:ab,ti

#26. #1-#25/ OR

#27. ‘glucocorticoid’/exp

#28. ‘methylprednisolone’/exp

#29. ‘cortisone’/exp

#30. ‘dexamethasone’/exp

#31. ‘prednisone’/exp

#32. ‘betamethasone’/exp

#33. ‘glucocorticoid*’:ab,ti

#34. ‘methylprednisolone*’:ab,ti

#35. ‘cortisone*’:ab,ti

#36. ‘dexamethasone*’:ab,ti

#37. ‘prednisone*’:ab,ti

#38. ‘budesonid*’:ab,ti

#39. ‘hexadecadrol’:ab,ti

#40. ‘cortodoxone’:ab,ti

#41. ‘hydrocortisone’:ab,ti

#42. ‘hydroxycorticosteroids’:ab,ti

#43. ‘adrenal cortex hormone*’:ab,ti

#44. ‘becl?met*’:ab,ti

#45. ‘betamet?asone*’:ab,ti

#46. ‘clobetasol*’:ab,ti

#47. ‘corticoid*’:ab,ti

#48. ‘corticosteroid*’:ab,ti

#49. ‘corticosterone*’:ab,ti

#50. ‘cortodoxone*’:ab,ti

#51. ‘dexamet?asone*’:ab,ti

#52. ‘glucocortico*’:ab,ti

#53. ‘hydrocortisone*’:ab,ti

#54. ‘hydroxycorticosteroid*’:ab,ti

#55. ‘hydroxypregnenolone*’:ab,ti

#56. ‘prednisolone*’:ab,ti

#57. ‘pregnenedione*’:ab,ti

#58. ‘pregnenolone*’:ab,ti

#59. ‘tetrahydrocortisol*’:ab,ti

#60. ‘triamcinolone*’:ab,ti

#61. #27-#60/ OR

#62. #25 AND #61

#63.#62 lim(embase)

#### Cochrane

#1. MeSH descriptor: (Middle East Respiratory Syndrome Coronavirus) explode all trees

#2. MeSH descriptor: (Severe Acute Respiratory Syndrome) explode all trees

#3. MeSH descriptor: (SARS Virus) explode all trees

#4. “COVID-19”:ti,ab,kw

#5. “SARS-COV-2”:ti,ab,kw

#6. “Novel coronavirus”:ti,ab,kw

#7. “2019-novel coronavirus” :ti,ab,kw

#8. “Novel CoV” :ti,ab,kw

#9. “2019-nCoV” :ti,ab,kw

#10. “2019-CoV” :ti,ab,kw

#11. “coronavirus disease-19” :ti,ab,kw

#12. “coronavirus disease 2019” :ti,ab,kw

#13. “COVID 19” :ti,ab,kw

#14. “Wuhan-Cov” :ti,ab,kw

#15. “Wuhan Coronavirus” :ti,ab,kw

#16. “Wuhan seafood market pneumonia virus” :ti,ab,kw

#17. “Middle East Respiratory Syndrome” :ti,ab,kw

#18. “MERS”:ti,ab,kw

#19. “MERS-CoV”:ti,ab,kw

#20. “Severe Acute Respiratory Syndrome”:ti,ab,kw

#21. “SARS” :ti,ab,kw

#22. “SARS-CoV” :ti,ab,kw

#23. “SARS-Related”:ti,ab,kw

#24. “SARS-Associated”:ti,ab,kw

#25. #1-#24/ OR

#26. MeSH descriptor: (Glucocorticoids) explode all trees

#27. MeSH descriptor: (Methylprednisolone) explode all trees

#28. MeSH descriptor: (Cortisone) explode all trees

#29. MeSH descriptor: (Dexamethasone) explode all trees

#30. MeSH descriptor: (Prednisone) explode all trees

#31. MeSH descriptor: (Budesonide) explode all trees

#32. MeSH descriptor: (Betamethasone) explode all trees

#33. “Adrenal Cortex Hormones” :ti,ab,kw

#34. “Glucocorticoid*” :ti,ab,kw

#35. “methylprednisolone*” :ti,ab,kw

#36. “cortisone*” :ti,ab,kw

#37. “Dexamethasone*” :ti,ab,kw

#38. “prednisone*” :ti,ab,kw

#39. “budesonid*” :ti,ab,kw

#40. “Beclomethasone” :ti,ab,kw

#41. “hexadecadrol” :ti,ab,kw

#42. “adrenal cortex hormone*” :ti,ab,kw

#43. “becl?met*” :ti,ab,kw

#44. “betamet?asone*” :ti,ab,kw

#45. “clobetasol*” :ti,ab,kw

#46. “corticoid*” :ti,ab,kw

#47. “corticosteroid*” :ti,ab,kw

#48. “corticosterone*” :ti,ab,kw

#49. “cortodoxone*” :ti,ab,kw

#50. “dexamet?asone*” :ti,ab,kw

#51. “glucocortico*” :ti,ab,kw

#52. “hydrocortisone*” :ti,ab,kw

#53. “hydroxycorticosteroid*” :ti,ab,kw

#54. “hydroxypregnenolone*” :ti,ab,kw

#55. “prednisolone*” :ti,ab,kw

#56. “pregnenedione*” :ti,ab,kw

#57. “pregnenolone*” :ti,ab,kw

#58. “tetrahydrocortisol*” :ti,ab,kw

#59. “triamcinolone*” :ti,ab,kw

#60. #26-#59/ OR

#61. #25 AND#60

#### Web of Science

#1 TOPIC: “COVID-19”

#2 TOPIC: “SARS-COV-2”

#3 TOPIC: “Novel coronavirus”

#4 TOPIC: “2019-novel coronavirus”

#5 TOPIC: “coronavirus disease-19”

#6 TOPIC: “coronavirus disease 2019”

#7 TOPIC: “COVID 19”

#8 TOPIC: “Novel CoV”

#9 TOPIC: “2019-nCoV”

#10 TOPIC: “2019-CoV”

#11 TOPIC: “Wuhan-Cov”

#12 TOPIC: “Wuhan Coronavirus”

#13 TOPIC: “Wuhan seafood market pneumonia virus”

#14 TOPIC: “Middle East Respiratory Syndrome”

#15 TOPIC: “MERS”

#16 TOPIC: “MERS-CoV”

#17 TOPIC: “Severe Acute Respiratory Syndrome”

#18 TOPIC: “SARS”

#19 TOPIC: “SARS-CoV”

#20 TOPIC: “SARS-Related”

#21 TOPIC: “SARS-Associated”

#22 #1-#21/ OR

#23 TOPIC: “methylprednisolone*”

#24 TOPIC: “cortisone*”

#25 TOPIC:”Dexamethasone*”

#26 TOPIC: “prednisone*”

#27 TOPIC: “budesonid*”

#28 TOPIC: “Beclomethasone”

#29 TOPIC: “hexadecadrol*”

#30 TOPIC: “adrenal cortex hormone*”

#31 TOPIC: “becl?met*”

#32 TOPIC: “betamet?asone*”

#33 TOPIC: “clobetasol*”

#34 TOPIC: “corticoid*”

#35 TOPIC: “corticosteroid*”

#36 TOPIC: “corticosterone* “

#37 TOPIC: “cortodoxone*”

#38 TOPIC: “dexamet?asone*”

#39 TOPIC: “glucocortico*”

#40 TOPIC: “hydrocortisone*”

#41 TOPIC: “hydroxycorticosteroid*”

#42 TOPIC: “hydroxypregnenolone* “

#43 TOPIC: “prednisolone*”

#44 TOPIC:”pregnenedione* “

#45 TOPIC:”pregnenolone*”

#46 TOPIC: “tetrahydrocortisol*”

#47 TOPIC: “triamcinolone*”

#48 #23-#47 OR

#49 #22 AND#48

#### CBM

#1. “新型冠状病毒”(常用字段:智能)

#2. “COVID-19”(常用字段:智能)

#3. “COVID 19”(常用字段:智能)

#4. “2019-nCoV”(常用字段:智能)

#5. “2019-CoV”(常用字段:智能)

#6. “SARS-CoV-2”(常用字段:智能)

#7. “武汉冠状病毒”(常用字段:智能)

#8. “中东呼吸综合征冠状病毒&#x201D;(不加权:扩展)

#9. “中东呼吸综合征&#x201D;(常用字段:智能)

#10. “MERS”(常用字段:智能)

#11. “MERS-CoV”(常用字段:智能)

#12. “严重急性呼吸综合征&#x201D;(不加权:扩展)

#13. “SARS 病毒”(不加权:扩展)

#14. “严重急性呼吸综合征”(常用字段:智能)

#15. “SARS”(常用字段:智能)

#16. #1-#15/ OR

#17. “糖皮质激素”(不加权:扩展)

#18. “糖皮质激素”(常用字段:智能)

#19. “可的松” (常用字段:智能)

#20. “布地奈德”(常用字段:智能)

#21. “地塞米松”(常用字段:智能)

#22. “强的松”(常用字段:智能)

#23. “泼尼松”(常用字段:智能)

#24. “甲泼尼龙”(常用字段:智能)

#25. “甲强龙”(常用字段:智能)

#26. #17-#25/ OR

#27. #16 AND#26

#### WanFang

#1. “新型冠状病毒”(主题)

#2. “COVID-19”(主题)

#3. “COVID 19”(主题)

#4. “2019-nCoV”(主题)

#5. “2019-CoV”(主题)

#6. “SARS-CoV-2”(主题)

#7. “武汉冠状病毒”(主题)

#8. “中东呼吸综合征”(主题)

#9. “MERS”(主题)

#10. “MERS-CoV”(主题)

#11. “严重急性呼吸综合征”(主题)

#12. “SARS”(主题)

#13. #1-#12/ OR

#14. “糖皮质激素”(主题)

#15. “泼尼松”(主题)

#16. “强的松”(主题)

#17. “布地奈德”(主题)

#18. “地塞米松”(主题)

#19. “可的松”(主题)

#20. “甲泼尼龙”(主题)

#21. “甲强龙”(主题)

#22. #14-#21/ OR

#23. #13 AND#22

#### CNKI

#1. “新型冠状病毒”(主题)

#2. “COVID-19”(主题)

#3. “COVID 19”(主题)

#4. “2019-nCoV”(主题)

#5. “2019-CoV”(主题)

#6. “SARS-CoV-2”(主题)

#7. “武汉冠状病毒”(主题)

#8. “中东呼吸综合征”(主题)

#9. “MERS”(主题)

#10. “MERS-CoV”(主题)

#11. “严重急性呼吸综合征”(主题)

#12. “SARS”(主题)

#13. #1-#12/ OR

#14. “糖皮质激素”(主题)

#15. “泼尼松”(主题)

#16. “强的松”(主题)

#17. “布地奈德”(主题)

#18. “地塞米松”(主题)

#19. “可的松”(主题)

#20. “甲泼尼龙”(主题)

#21. “甲强龙”(主题)

#22. #14-#21/ OR#23.

#13 AND#22

## Notes

### Competing Interest Statement

The authors have declared no competing interest.

